# Detecting simulated pathogen releases in a real-world health data set

**DOI:** 10.64898/2026.05.12.26350999

**Authors:** Robert Moss, Matthew J. Testolin, Christos Pitsaris, Alexander M. Hill, David J. Muscatello, James M. McCaw, Peter Dawson

## Abstract

The purpose of electronic disease syndromic surveillance (EDSyS) systems is to detect hazardous pathogens and other unusual signals in health surveillance data before such events are identified by an individual clinician or healthcare facility. However, EDSyS systems have primarily been evaluated using simulated health surveillance data, which do not necessarily capture the richness and complexities of real-world health data. We have updated and extended an existing EDSyS system, EpiDefend, which combines ensemble forecasting and recursive Bayesian estimation in a particle filter framework that supports demographic and spatial structure. We simulated the release of several pathogens, both infectious and non-infectious, and injected the resulting cases into a real-world health data set. Here we evaluate EpiDefend’s sensitivity and specificity in detecting these simulated releases, and measure the time to detection against pathogen-specific estimates of the time to clinical detection, as informed by clinicians and microbiologists. We show that for diseases where clinical diagnosis can be challenging, such as Q fever (Coxiella burnetii) and tularaemia (Francisella tularensis), EpiDefend can reliably beat the time to clinical detection. In contrast, for pathogens that can be clinically diagnosed relatively quickly, such as inhalational anthrax and pneumonic plague, it is extremely difficult to beat the time to clinical detection. Our results suggest that EpiDefend may be able to reliably detect real-world introductions or releases of some pathogens at low false-alarm rates before a clinical diagnosis would be confirmed, and this would represent a landmark achievement for EDSyS systems.

## 1 Introduction

The release or spread of a hazardous pathogen can rapidly disrupt health services and cause substantial morbidity and mortality, as most recently demonstrated by the COVID-19 pandemic [1]. Emergency preparedness and response capabilities are crucial for protecting health systems and populations from emerging disease threats [2] and deliberate releases (bioterrorism) [3]. Electronic disease syndromic surveillance systems (EDSyS, also known as early warning systems) are designed to rapidly detect unusual signals in syndromic health surveillance data, so that response actions can be taken in a timely manner [4]. An EDSyS can potentially identify a release more quickly than an individual clinician, clinic, or health facility, by analysing patient data at larger scales, such as an entire health system.

However, while a recent review by Patel et al. [5] of 29 systems confirmed that EDSyS has a critical role in “enhancing public health responses to respiratory infections”, the authors also identified a need for continued innovation, with common limitations including a lack of generalisability, selection bias, and small sample size for model validation. An earlier review of Emergency Department syndromic surveillance systems by Hughes et al. [4] identified similar limitations, and flagged the potential for collaborative improvements by sharing indicator details and harmonisation between systems. Both reviews highlighted the challenge of using diagnosis codes as indicators, given their the limited accuracy for identifying specific pathogens or syndromes of concern [6–9].

Here, we present a methodological extension of the “EpiDefend” EDSyS [10] and evaluate its performance in detecting simulated releases of several pathogens embedded in retrospective real-world health surveillance data from the PEARL real-world linked health database [11–15]. We used a variety of evidence sources (including published articles, laboratory guidelines, and conversations with clinicians and pathologists) to estimate the time at which each simulated release would likely be detected by a clinician, and used these detection times as the benchmark for evaluating EpiDefend’s detection performance.

By evaluating the EpiDefend’s performance using real-world health data and for a variety of pathogens (both infectious and non-infectious) with distinct characteristics, we address the common limitations of existing EDSyS systems identified by Patel et al. [5]. We outline how the methods presented here can be generalised to capture greater details in population and spatial structure, and conclude that real-world evaluation of EpiDefend is a plausible next step.

## 2 Materials and Methods

The simulated release scenarios, input health data, and EpiDefend EDSyS algorithm involve a number of distinct components, as outlined in Figure 1.

**Figure 1:**
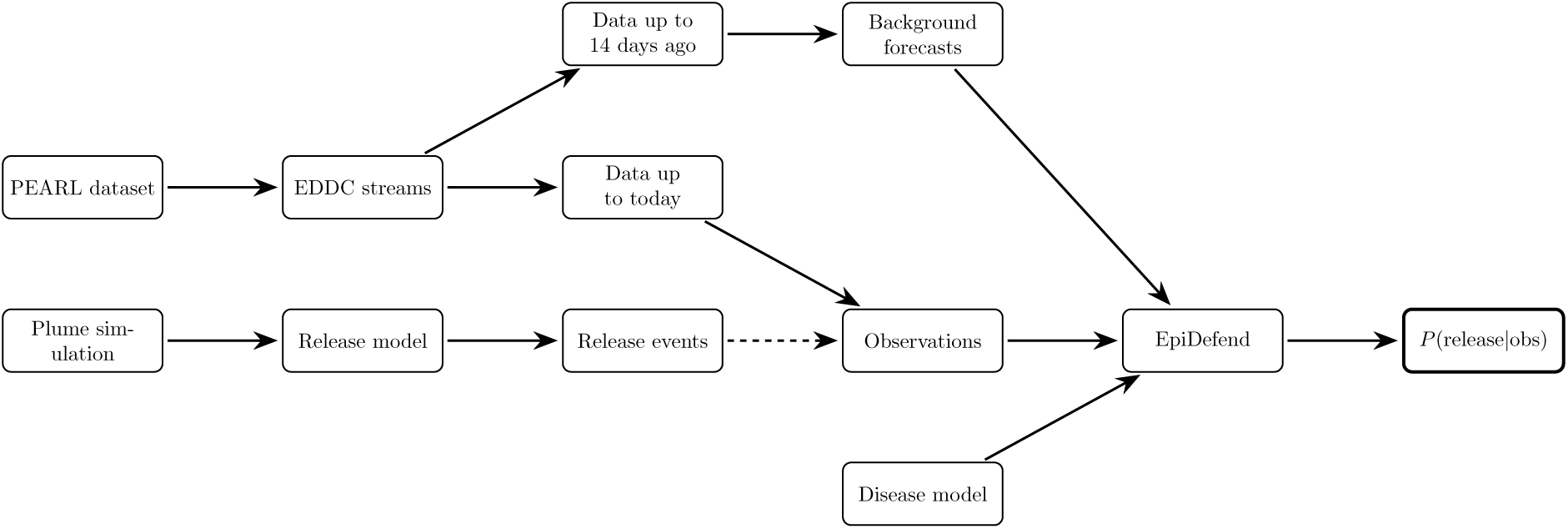
An overview of the data sources, release simulations, and inference method. Note that healthcare events arising from a simulated release may or may not be included in the input observations (indicated by the dashed arrow). The output is an estimated probability that cases arising from a release are present in the input observations.

In brief, we used retrospective emergency department arrivals and critical care admissions data (“EDDC”) from the PEARL health database to characterise normal (“background”) healthcare activity for the EpiDefend inference algorithm.

Then, for each pathogen, we used a high-resolution simulation model (“plume simulation”) to generate spatial pathogen concentration profiles, and combined this with population demographic and mobility data (“release model”) to simulate exposures to the pathogen, the progression of symptoms, and seeking healthcare. These synthetic healthcare events (“release events”) were then injected into to the PEARL data streams (“data up to today”) to form the input health data for EpiDefend (“observations”).

The EpiDefend inference algorithm (“EpiDefend”) estimates the probability of a release occurring in the past 14 days, *P* (release|obs), by combining statistical forecasts of the normal healthcare activity over this period (“Background forecasts”), the healthcare events observed over the past 14 days (“observations”), and a pathogen-specific model of plausible release events (“disease model”). To evaluate both true-detection and false-detection rates, we included scenarios where the input health data (“observations”) did not contain a simulated release.

We now describe the PEARL data streams and each modelling component in turn.

### 2.1 Ethics statement

The New South Wales Population Health and Health Services Research Ethics Committee granted ethics approval for this study (2021/ETH00070).

### 2.2 Health data

We used the line-listed Emergency Department Data Collection (EDDC) data set, which defines the patient cohort included in the PEARL database and captures emergency department presentations, to characterise healthcare events in which we would expect to observe cases arising from a biological release. Given the diversity of diagnosis codes that can be assigned to the same underlying pathogen [6–9], we only included presentations that met one or more predefined syndromic definitions associated with long-term ICU episodes caused by influenza and COVID-19 infections [12, 15], and which occurred between 2013 and 2018 (inclusive).

We used daily counts of syndromic presentations for five different age group — 0-4 years, 5-18 years, 19-65 years, 66+ years, and all ages. We also used daily counts of “Critical admission”events, which we defined as syndromic patients who arrive by ambulance (according to the “ED mode of arrival” field) and are admitted directly into critical care (according to the “ED discharge status” field). These are the six PEARL data streams into which we injected synthetic healthcare events. By aggregating events across metropolitan Sydney, rather than having separate data streams for each facility, we reduced the signal to noise ratio in these data streams, and simplified the inference problem. The rationale for separate age groups is that the age distribution of cases arising from a release event is likely to differ from the business-as-usual age distribution of syndromic presentations, and so these age-specific data streams may provide additional information and improve the sensitivity of the EpiDefend inference algorithm.

### 2.3 Characterise normal health signals

The aim of EpiDefend is to estimate the probability that, among all cases reported in the health data that satisfy the syndromic criteria for a possible infection by a specific pathogen of interest over the past 14 days, at least one of these cases is due to a release of that specific pathogen. We assumed that health data before this 14-day period do not include any cases arising from a release, on the grounds that such releases would already have been detected via clinical diagnosis or standard surveillance systems. Accordingly, we treated the health data for the past 14 days as suspect, and used an equal-weights ensemble of regression models to produce probabilistic forecasts for each data streams (see subsection A.1) based on the health data up to 14 days ago. We generated these forecasts for every 14-day window in the study period.

### 2.4 Chosen pathogens

We selected four bacterial pathogens from the Australian Security Sensitive Biological Agents list [16] that are plausible bioterrorism candidates and which result in symptoms consistent with common respiratory infections: inhalational anthrax, pneumonic plague, Q fever, and tularaemia. For each pathogen, we characterised the progression from initial exposure to the onset of severe disease (for which we assume that infected persons will rapidly seek healthcare). For anthrax, we also characterised an interim period of mild (“prodromal”) symptoms, during which an infected person may also seek healthcare. We characterised the residence time for each period using log-normal distributions (Table 1).

**Table 1:** Log-normal distributions for the incubation per each pathogen, and the prodromal period for anthrax, with residence times *T* ∼ Lognormal(exp(*M*), *σ*). See subsection A.2 for further details.

| Pathogen | Period | Median (M) | Log-sd ($\sigma$) |
| --- | --- | --- | --- |
| Anthrax | Incubation | 5 days | 0.4 |
|  | Prodromal | 2.3 days | 0.3 |
| Pneumonic plague | Incubation | 4 days | 0.4 |
| Q fever | Incubation | 18 days | 0.343 |
| Tularaemia | Incubation | 4 | 0.76 |

### 2.5 Release simulations

Release events were simulated using meteorological conditions typical of mid-August in Sydney. Accordingly, our study period spans the months of July to September (inclusive), which comprise a reasonable window over which to train and evaluate the EpiDefend algorithm. For each pathogen we used the Hazard Prediction and Assessment Capability (HPAC) model to simulate releases for each combination of source mass size, release location, and meteorological conditions for different times of day [17–19]. Six release locations were chosen across metropolitan Sydney, including the central business district, sports facilities and schools, residential suburbs, tourist destinations, industrial districts, and major commercial centres. The locations differed markedly in their residential populations and work trips, as shown in Table 2 (see subsection A.3 for further details).

**Table 2:** Resident population and daily work trip counts for each release location. People who reside outside of the release location and travel there for work are counted in the “Trips in” column, while people who reside in the release location and travel elsewhere for work are counted in the “Trips out” column.

| Location | SA2 | Population | Trips in | Trips out |
| --- | --- | --- | --- | --- |
| Martin Place | 117031337 | 29,636 | 317,675 | 14,503 |
| Moore Park | 118011345 | 16,280 | 9,333 | 8,623 |
| Balmain | 120021387 | 16,629 | 4,844 | 8,031 |
| Manly | 122011419 | 23,655 | 8,602 | 11,832 |
| Banksmeadow | 117011320 | 21 | 5,572 | 9 |
| Parramatta | 125041492 | 30,480 | 49,934 | 13,954 |

The release masses were calibrated so that in a reference release simulation, the plume of each pathogen would infect 10, 50, 250, and 1250 people, for the small, medium, large and huge scenarios, respectively. These reference masses were then used in the simulations for each of the six release locations. The simulated releases for this study typically resulted in significantly larger numbers of infections than the reference release simulations, due to the population density and large trip volumes in each release location.

**Figure 2:**
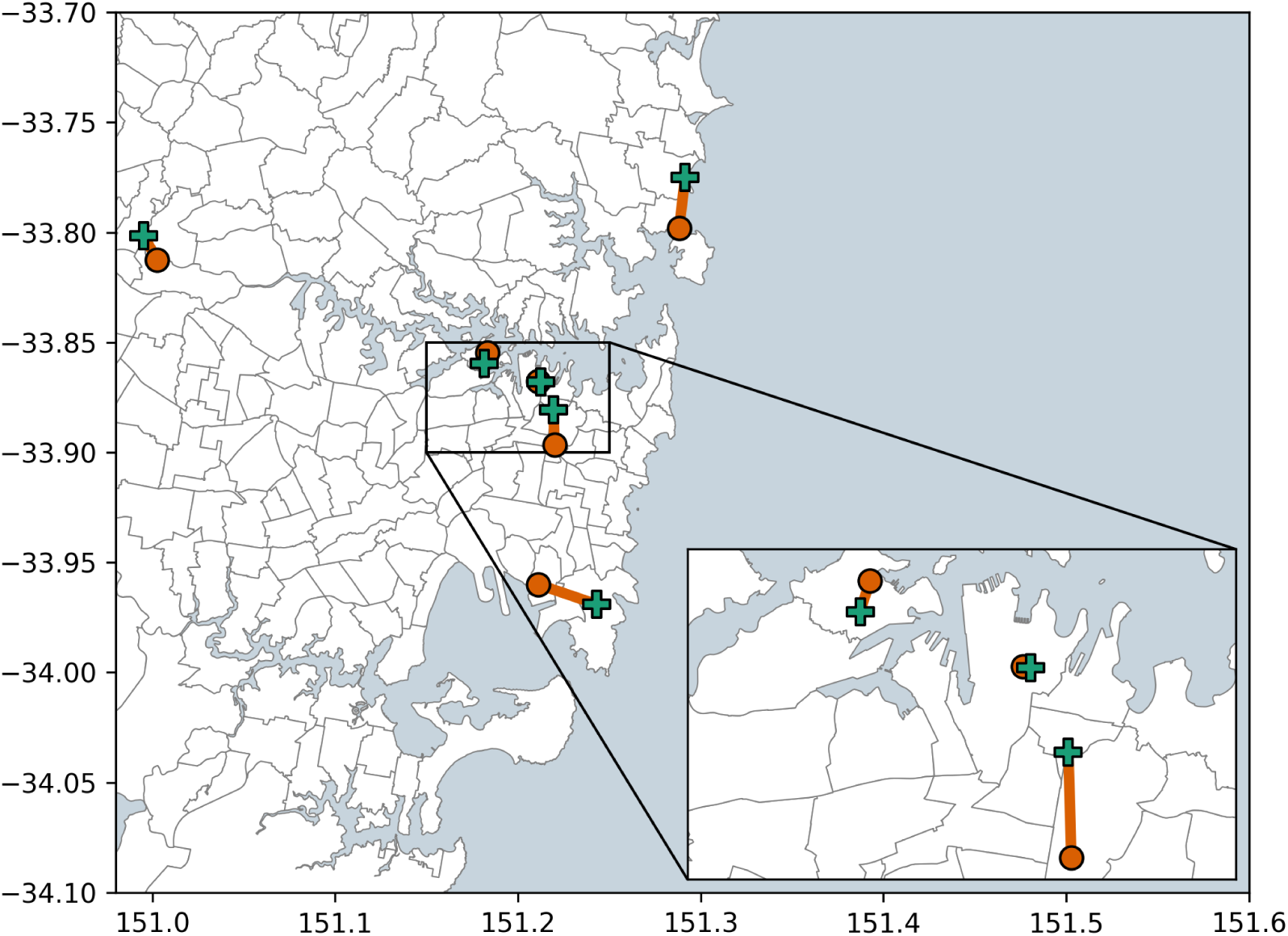
A map of the plume release locations (red circles). Outer locations, clockwise from top-right: Manly, Banksmeadow, Paramatta. Inner locations (inset), top-left to bottom-right: Balmain, Martin Place, Moore Park. Red lines connect each release location to the nearest hospital (green crosses). Dark lines show SA2 boundaries.

### 2.6 Synthetic healthcare events

The outputs of each plume simulation were time-series of pathogen concentrations *c*(*x*_*i*_, *y*_*i*_, *t*) for a spatial grid with samplers at locations (*x*_*i*_, *y*_*i*_) ∶ *i* ∈ {1, 2, …, 2653} over 30 minutes (*t* ∈ {1, 2, …, 1800} seconds).

Each individual *p* located in any SA2 in this spatial grid had a time-varying location {*L_t_*^*p*^} and was exposed to a net dose *d_p_*. From this net dose we calculated the probability of that individual being infected *P* (*E*|*d*_*p*_) using a North Atlantic Treaty Organization (NATO) standard probit model for dose response [20]. We followed Haber’s rule and assumed that this probability depends only on the net dose and not the exposure duration [21], treated each individual’s outcome as a binomial process, and calculated the number of exposures *E*_*N*_ in each SA2:

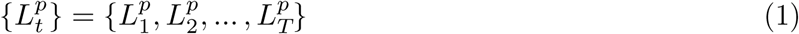

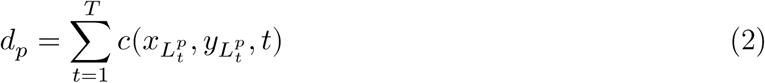

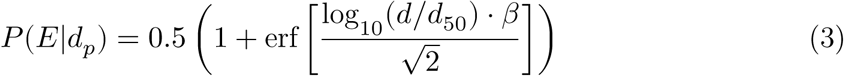

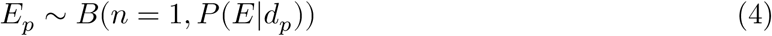

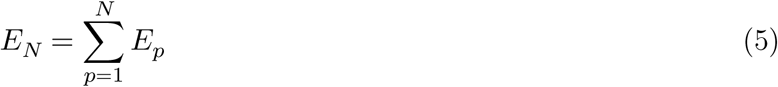

For individuals located indoors, the reduction in exposure can be estimated from a simple compartment model via a protection factor (PF), based on typical building air exchange rates and exposure duration [20]. When the duration of external exposure is brief, as per the release simulations used here, and building occupancy times are long (e.g., up to 8 hours for a commercial building, potentially even longer for residential buildings), then *P F* ≈ 1 and the reduction in exposure is negligible. Accordingly, we assumed that the probability of exposure for a given dose did not depend on whether the individual was located outdoors or indoors.

To characterise the movement of exposed individuals between SA2s, we used Australian Bureau of Statistics (ABS) Journey to Work (JTW) data to define the daily journeys *J*(*A*_*i*_, *A*_*j*_) from SA2 *A*_*i*_ to SA2 *A*_*j*_, and the ABS estimated resident populations *N*_*R*_(*A*), to calculate the working populations *N*_*W*_(*A*) and movement probabilities for travelling to work (9) and returning home (10):

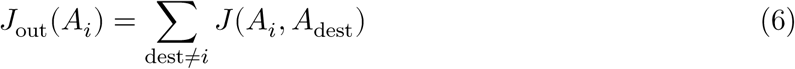

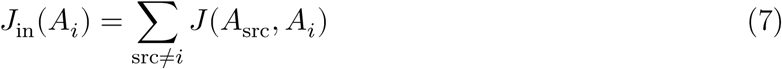

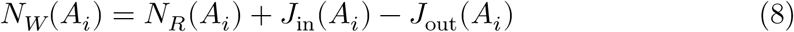

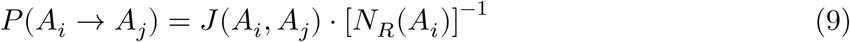

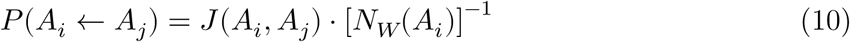

For an individual exposed to the pathogen in SA2 *A*_*E*_ at time *t*_*E*_, we defined the probability *P* (*A*_*i*_|*A*_*E*_, *t*) of that individual being located in SA2 *A*_*i*_ at time *t* in terms of these movement probabilities, under the assumption that individuals are at work during times {*t*_*W*_} and are at home during all other times {*t*_*R*_}:

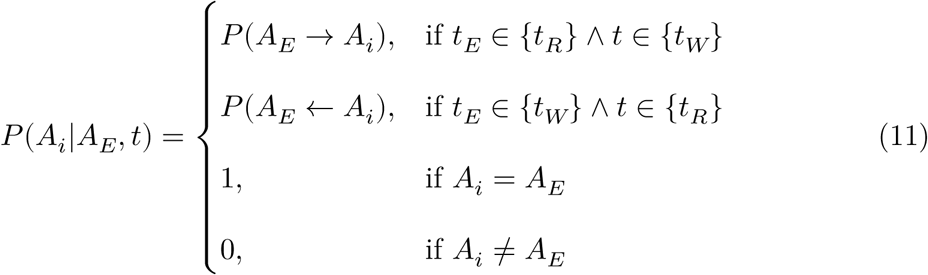

When an individual’s symptom progression resulted in a healthcare-seeking event, we sampled their current SA2 *A*_*i*_ (11) and selected the healthcare facility 𝓗_*h*_ ∈ 𝓗 nearest to the centroid *C*_*i*_ of *A*_*i*_, using a data set of Australian hospital locations [22]:

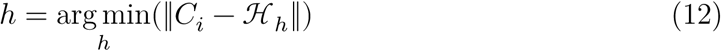

An individual’s symptoms and severity for each healthcare-seeking event were sampled from pathogen-specific models of disease progression. For anthrax, pneumonic plague, and tularaemia, we assumed that all exposed individuals experience rapid onset of severe disease (fulminant stage) and will rapidly seek healthcare as a consequence. This ignores the possible provision of antibiotics to anthrax cases who present while in the prodromal stage and may avoid the fulminant stage. In contrast, for Q fever we assumed that 60% of exposed individuals never develop symptoms, that 38% of exposed individuals present with acute self-limited symptoms, and only 2% of exposed individuals experience rapid onset of severe disease (fulminant stage) and require hospitalisation [23]. We assumed that 50% of cases with fulminant disease will arrive via ambulance.

The delay from the onset of severe disease to arriving at an Emergency Department is not observable, and cannot be estimated from available data sets. Consistent with prior work, including previous iterations of the EpiDefend algorithm, we assumed that this delay followed a geometric distribution characterised by the hourly probability *p*_seek_ of seeking healthcare given severe disease. If we assume that most, but not all, people will arrive at a hospital within 12 hours of developing clinically severe symptoms, we find 0.1 ≤ *p*_seek_ ≤ 0.2.

To simulate human-to-human transmission of pneumonic plague, we modelled the number of secondary infections *E*^*p*^*_N_* for each exposed individual *p* as a Poisson process: *E*^*p*^*_N_* ∼ Pois(*λ* = *R*), and sampled the exposure times *t*_*E*_ from individual *p*’s infectious period using a uniform distribution. Exposure locations *A*_*E*_ were sampled according to individual *p*’s current location (11), and their subsequent movements and healthcare events were simulated as per the primary cases. For a given time horizon *T* after a release at time *t* = 0, we repeatedly simulated secondary infections until there were no exposures for which *t*_*E*_ < *T*.

### 2.7 Time to clinical detection

For each pathogen, we defined delay distributions to estimate the time at which a public health clinician would be likely to identify the release pathogen in a clinical case. These delay distributions are the benchmarks against which the detection performance of the EpiDefend algorithm was compared and evaluated. Each pathogen-specific model shares the same sequence of events that are necessary for a clinical detection to occur, and the differences between the pathogen-specific models are captured in the delay distributions for disease progression and laboratory testing (see Figure 3).

**Figure 3:**
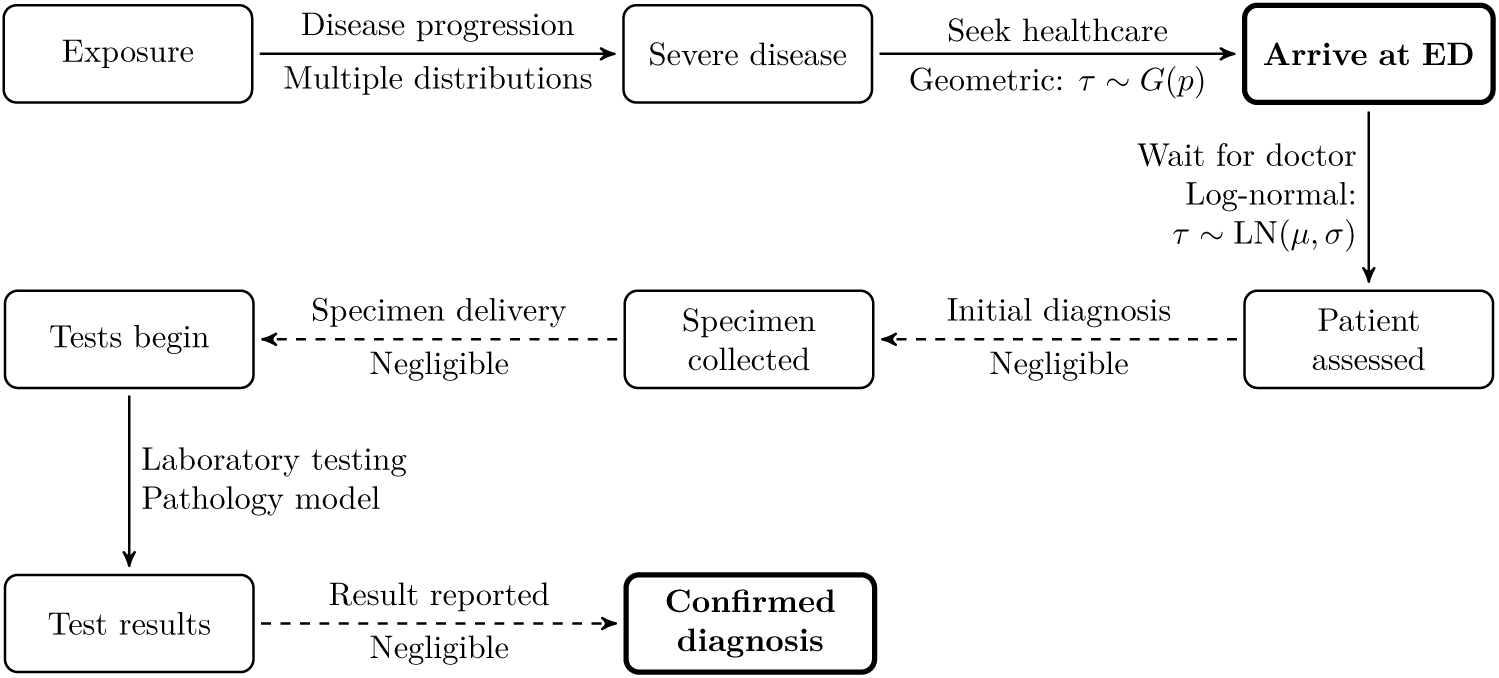
A diagram of the sequence of events that begins with an initial exposure and culminates in a clinical detection of the release. The time to clinical detection is the delay between arriving at the ED (bold node) and a confirmed diagnosis (bold node).

The delay between arriving at an Emergency Department and being assessed by a clinician was informed by a 2009 study of 501 pneumonia patients in an Australian emergency department, which reported the delay between the time of presentation to the hospital triage desk, and the documented time of first antibiotic administration in the medication chart [24]. We obtained a good fit to these delay times *τ* with a log-normal distribution (see subsection A.4 for further details):

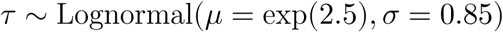

Estimated laboratory testing delays are shown in Table 3. While Q fever can be detected by RT-PCR, this is an unlikely diagnosis method in an urban setting. For tularaemia, there is currently no serology testing in Australia, RT-PCR testing is limited, and routine blood cultures will not return a positive result. Accordingly, urban mass-exposure events for Q fever and Tularaemia would not be rapidly detected by standard pathology testing. Metagenomic approaches would be used to identify the pathogen(s) responsible for a sufficiently large spike in unexplained fevers, but we do not know what level unknown fevers would trigger this response in a given hospital.

**Table 3:** Estimated laboratory testing delays for each pathogen. ^∗^We chose best-case delays instead of these long delays, as described in the main text.

| Pathogen | Diagnosis method | Delay |
| --- | --- | --- |
| Anthrax | MALDI-TOF | 15–19 hours |
| Pneumonic plague | MALDI-TOF | 15–19 hours |
| Q fever | Serology | 7–10 days* |
| Tularaemia | Blood culture | 7–10 days* |

To avoid evaluating EpiDefend’s performance against such long diagnosis delays for Q fever and tularaemia, where success is all but guaranteed, we have instead assumed best-case diagnosis delays for these pathogens. For Q fever we chose a testing delay of 48 hours, assuming positive RT-PCR results within several days of symptom onset. For tularaemia we chose a testing delay of 72 hours, assuming that the pathology laboratory uses MALDI-TOF mass spectrometry [25, 26] and that the MALDI-TOF library contains a Francisella entity [27].

### 2.8 EpiDefend disease models

To simulate exposed individuals’ disease progression and healthcare events in EpiDefend (the “disease model” in Figure 1) we used a stochastic SEIR model. The *E* and *I* compartments were used to represent the incubation and prodromal stages of disease, and the *R* compartment represented the end of the prodromal stage (i.e., either the onset of fulminant disease, or recovery from infection for pathogens where this is a possible outcome). The number of *E* and *I* compartments, and their associated rate parameters (*σ* and 𝛾), were different for each pathogen, to approximate the incubation and prodromal periods listed in Table 1. We used a single *I* compartment to represent the final pre-fulminant stage for pneumonic plague, Q fever, and tularaemia (since they do not include a prodromal stage).

The disease model state vector comprised the time of the release *τ*_0_, the number of initial exposures *e*_0_, the SEIR model parameters and state variables, and a background forecast trajectory index *f*:

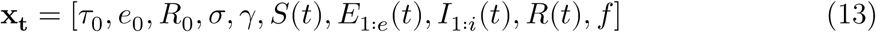

Of the four pathogens, only pneumonic plague allows for human-to-human transmission; for the other pathogens, we assumed *R*_0_ = 0 (see subsection A.5).

### 2.9 Sequential Monte Carlo inference

We used the post-regularisation particle filter (PRPF) to simulate many realisations of the disease model (“particles”) and update particle weights at each time-step to reflect how well each particle explained the observed data streams. For each 14-day window in the study period (using a 1-day slide interval), and each of the simulated pathogen releases, we generated 15 data sets:

- One data set using only the ground-truth PEARL data (i.e., no release); and
- Fourteen data sets with the simulated release cases added to the PEARL data, one data set for each day in the window.

The first data set allows us to measure EpiDefend’s false-alarm rate over the entire study period, while the other data sets allow us to measure EpiDefend’s true-alarm rate for all release dates and lead times in the study period.

For each data set, the estimated probability of a release was recorded at each day of the window (see below). Additionally, for each data stream, we recorded forecast credible intervals for (a) the entire particle ensemble; (b) only particles with a least one infection that may have resulted in an observed case; and (c) only particles with no infections that may have resulted in an observed case. The distinction between (b) and (c) is important, because until an infection reaches a disease state that can result in an observable healthcare event, the infection is invisible (i.e., not present in any of the data streams). Accordingly, particles for which there are only invisible infections do not contribute to the estimated probability of a release.

Because Q fever has a substantially longer incubation period than the other pathogens, we shifted the simulated attack cases backwards by one week and started the estimation process one week earlier, using the ground-truth PEARL data for the first 7 days of the 21-day window.

Our prior distributions for the data streams are the product of (a) the background forecast distribution for that data stream; and (b) the release cases distribution for that data stream. Since we are using an SMC framework, we approximate this by drawing a finite number of samples from both distributions. Accordingly, each particle comprises a state vector for the disease model and a sample trajectory from the background forecast for each data stream, and this allows for joint estimation of the background signal *f*_*t*_ and the release signal *c*_*t*_. If we instead marginalised over the background forecast distribution (with each particle comprising only a state vector for the disease model) we would be unable to account for bias in the background forecast and would obtain biased estimates of the probability of a release. To illustrate, if the background forecast systematically over-estimated the true background signal we would systematically underestimate the release probability, and if the background forecast systematically under-estimated the true background signal we would systematically overestimate the release probability.

Observable healthcare events arise from both prodromal and fulminant presentations. For anthrax infections, we assumed that prodromal presentations occurred at the midpoint of the prodromal stage (i.e., when an individual enters compartment *I*_6_), and we set the prodromal visibility index 𝑣 = 6. For all other pathogens, there is no prodromal stage and we set 𝑣 = *i* + 1. With this index 𝑣 we defined the subset *V* of all particles **x_t_** for which there has been at least one infection that may have resulted in an observed case (14), and sum the weights 𝑤_*i*_ of all particles in *V* to define the probability of a visible release at time *t* (15):

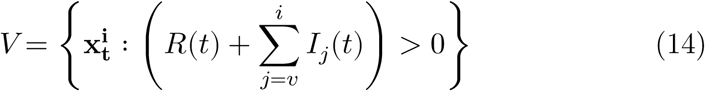

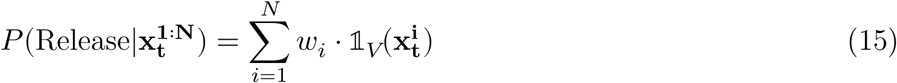

For age-specific and all-age respiratory EDDC presentations, the likelihood of observing **y_t_**cases was modelled as a negative binomial process with dispersion parameter 𝑘, where we have an expected value 𝐵_*f*_ from the background forecast trajectory *f*, we expect a proportion *p*_mild_ of the *N*_mild_ prodromal anthrax cases to present, and assume a fraction *A* of exposures occur in this age group:

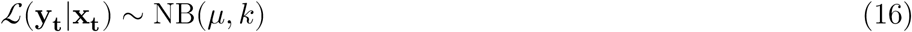

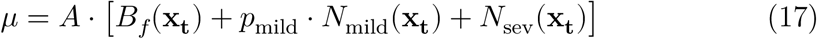

where the equations for *N*_mild_ and *N*_sev_ are defined in subsection A.6.

For anthrax infections, the number of mild presentations over the past day is simply the number of individuals who entered the mid-point compartment *I*_𝑣_ multiplied by the presentation fraction *p*_mild_. The number of severe presentations over the past day is more complex to calculate, due to the geometric delay distribution for the time between onset of fulminant disease (entering the *R* compartment) and arriving at a hospital. We consider individuals who entered *R* in each of the past 48 hours, and for each hourly window calculate the probability that these individuals would have arrived at a hospital in the past 24 hours.

For ambulance arrivals that are directly admitted to critical care, the likelihood of observing **y_t_** cases was modelled as a binomial process for a total population size *N* and with *N*_sev_ individuals having experienced the onset of severe symptoms, of which we expect a proportion *p*_amb_ to arrive by ambulance:

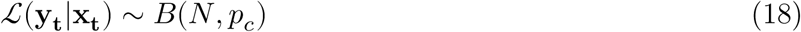

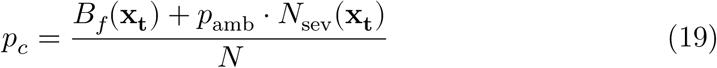

### 2.10 Performance evaluation

A detection alarm occurs at time *t* if the estimated probability of a release exceeds a given detection threshold *P*_*T*_ ∈ (0, 1]:

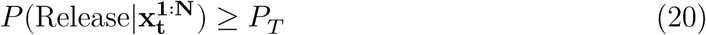

We evaluate EpiDefend’s detection performance over a range of detection thresholds *P*_*T*_ by quantifying the trade-off between detection timeliness and the false-positive rate with the use of Activity Monitor Operator Characteristic (AMOC) curves. Here we present AMOC curves using the standard timeliness measure of the delay between the initial release and detection, and a second set of AMOC curves that measure timeliness with respect to TTCD (i.e., the detection lead/lag time).

Once a detection alarm occurs, it is possible to investigate the “backcast” observation likelihoods ℒ(**y_1_**_∶**t**_|**x_t_**) and, in particular, to calculate these likelihoods separately for the subset of particles with a visible release (ℒ_release_) and the subset of particles without a visible release (ℒ_normal_), to identify the observation(s) that had the greatest influence on the estimated probability of attack:

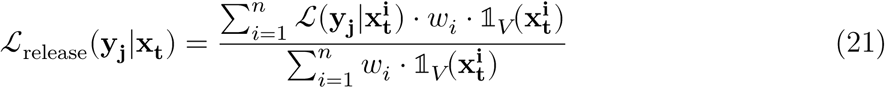

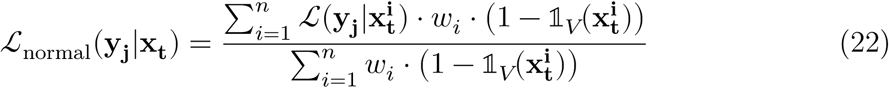

## 3 Results

### 3.1 An example release

For illustrative purposes, we first consider a single simulated anthrax release that occurs on day 9 of a 14-day window (i.e., 5 days before “today”, 12th September). We note here that critical admission counts and forecasts are not shown, due to data custodian privacy concerns regarding small counts. As shown in Figure 4, our background forecasts for this 14-day window are in good agreement with the true PEARL data (yellow circles, not made available to EpiDefend). The observations provided to EpiDefend (pink crosses) include healthcare events arising from the simulated release. The time of the release is indicated by the black vertical dashed line, and it takes several days before presentations due to this release become visible in the data.

**Figure 4:**
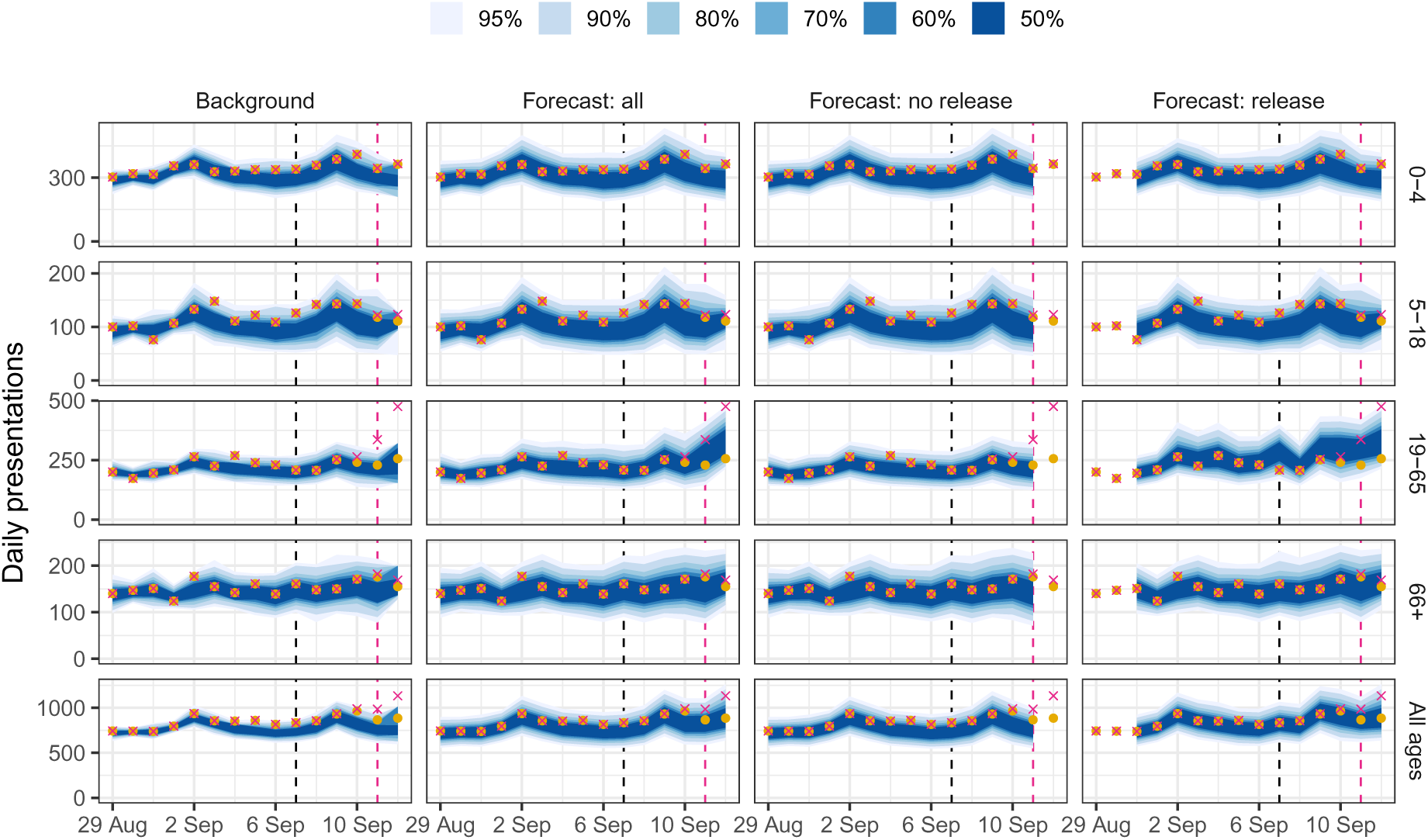
Example forecasts for a 14-day window which contains a simulated release, shown separately for each age group (rows). Columns show (a) the input background forecasts; (b) the EpiDefend backcasts; (c) the EpiDefend backcasts for particles without a visible release; and (d) EpiDefend backcasts for particles with a visible release. Forecast credible intervals (shaded blue regions) are plotted against the provided observations (pink crosses) and background PEARL data (yellow circles, not made available to EpiDefend). The black vertical dashed line indicates the date of the simulated release, and the pink vertical dashed line indicates the date when the estimated probability of a release first exceeds the threshold *P*_*T*_ = 0.5.

Four days after the release, EpiDefend estimates there is a greater than 50% probability of a release, as indicated by the pink vertical dashed line. Daily estimates of this probability over this 14-day window are shown in Figure 5. The probability of a release remained very low for the first three days after the release, then increased rapidly. Five days after the release (“today”) the estimated probability of a release was 1 (i.e., absolute certainty that there is a release). This means that every particle in the ensemble has a visible release, and so the “no release” forecast ends four days after the release (“yesterday”).

**Figure 5:**
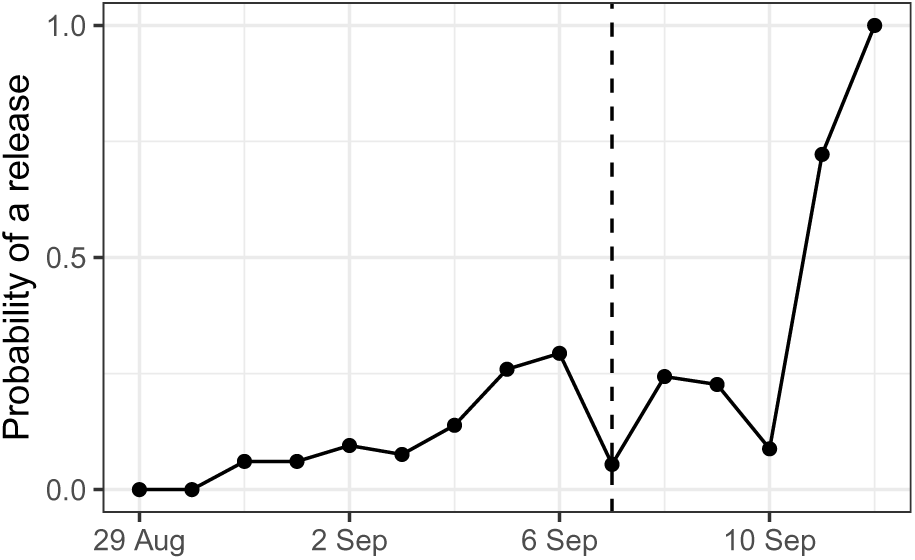
The estimated probability of a release for the simulation shown in Figure 4.

If we chose a detection threshold between 0.3 and 0.7, EpiDefend would signal an alarm 4 days after the simulated release. Recall that we have assumed a median delay of 2.5 hours for being assessed by a clinician and 15–19 hours for a confirmed anthrax diagnosis. The first severe presentation occurred 3 days after the simulated release, and so we expect this release to be identified by a clinician 4 days after the release. The outcome for this release is that EpiDefend’s time to detection is equivalent to that of a clinician. If we chose a detection threshold higher than 0.7, EpiDefend would only signal an alarm 5 days after the simulated release, 1 day later than the clinical detection.

### 3.2 EpiDefend sensitivity and specificity

We now consider EpiDefend’s performance for the medium-size release simulations for each pathogen. False alarm and release detection rates are shown in Figure 6 as a function of the detection threshold *P*_*T*_(20). Low false alarm rates can be achieved with detection thresholds of *P*_*T*_ ≈ 0.6 for medium releases of all pathogens except Q fever, for which the false alarm rate is much higher than the other pathogens for any given detection threshold because of the substantially smaller number of clinical events in the Q fever releases. This relationship between simulated attack case counts and false alarm rate is also evident in the different false alarm rates for small and medium anthrax releases.

**Figure 6:**
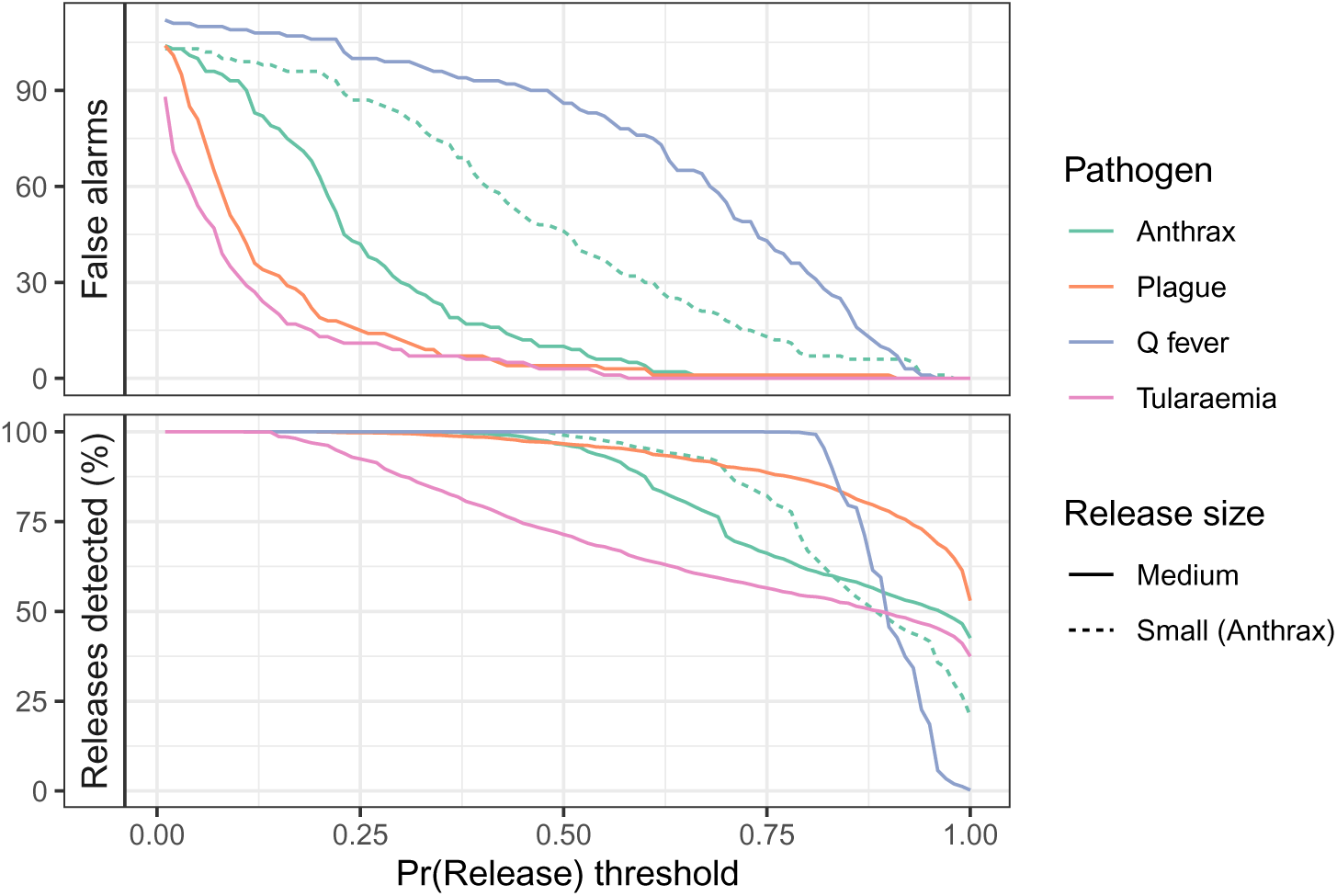
Top panel: The number of false alarms raised over the 2018 winter for each scenario. Bottom panel: The percentage of simulated attacks that were detected.

Detection sensitivity is also inversely proportional to the detection threshold *P*_*T*_(Figure 6). Plague and tularaemia have the lowest false alarm rates, but markedly different detection sensitivity. Detection sensitivity profiles for each release location clearly demonstrate the effect of population density and case numbers; all releases in the Sydney CBD (Martin Place) are detected, except for Q fever releases at the highest detection thresholds (see the supplementary materials). Detection sensitivity is lower for releases in the other locations, with the lowest sensitivity in Banksmeadow (extremely small resident population and second-fewest incoming work trips) and the highest sensitivity in Parramatta (largest resident population and second-most incoming work trips).

### 3.3 EpiDefend detection timeliness

We now consider the trade-off between detection timeliness and false alarm rates. The most meaningful benchmark for detection timeliness is the time to clinical detection (TTCD). Figure 7 (top panel) shows the relationship between the false alarm rate and the lead time of EpiDefend detections versus clinical detections, where positive values indicate that EpiDefend detected the attack earlier than the TTCD.

**Figure 7:**
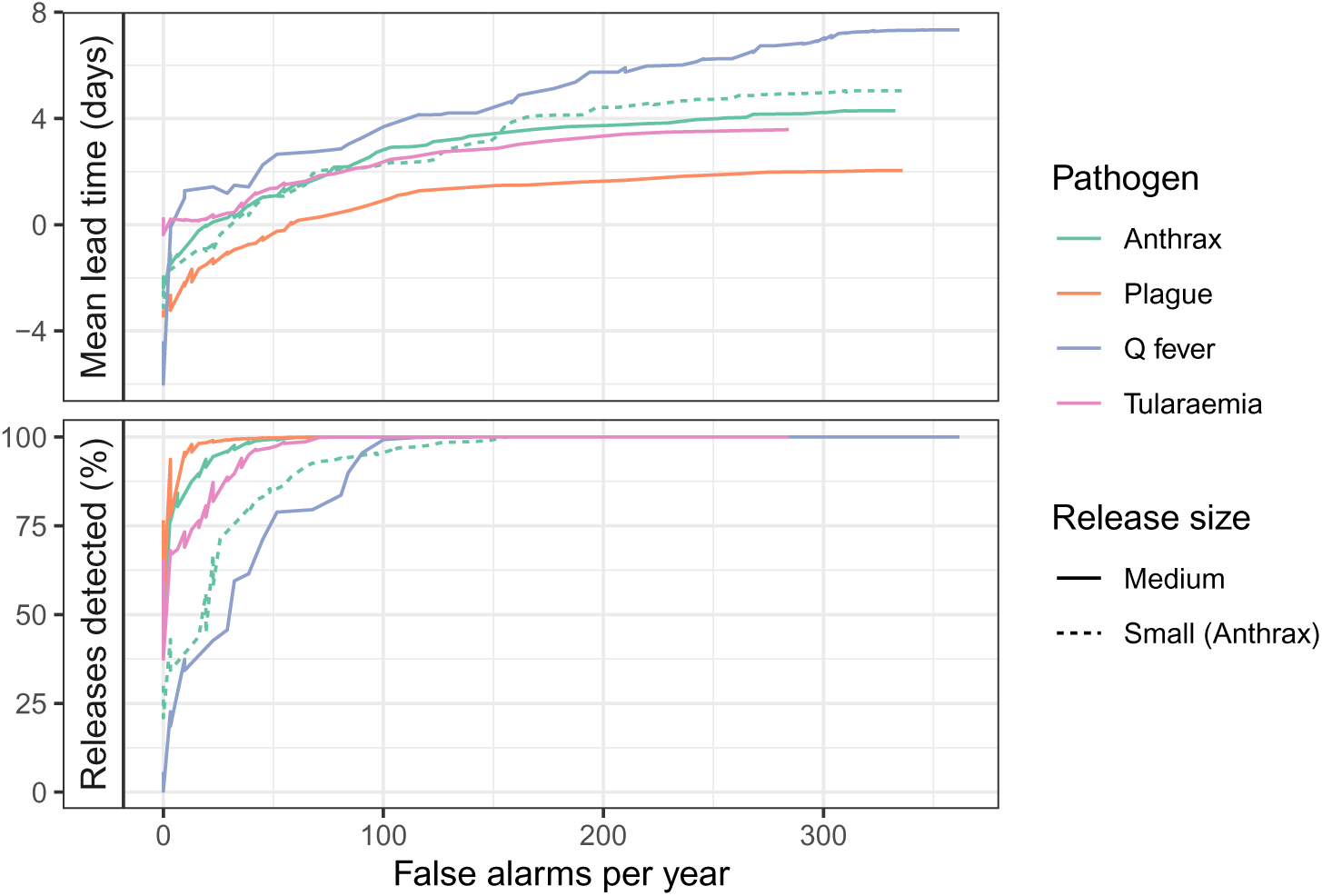
Top panel: The mean lead time relative to clinical detection, where positive values indicate that EpiDefend detected the attack before the clinical detection. Bottom panel: The percentage of attacks that were detected by EpiDefend.

For Q fever and tularaemia — where we have assumed best-case clinical diagnosis delays — EpiDefend is able to detect attacks as quickly as, or even quicker than, the TTCD at extremely low false alarm rates. For anthrax releases, EpiDefend can match the TTCD at slightly higher alarm rates, while pneumonic plague releases require the highest false alarm rates in order to match or beat the TTCD. Investigating the lead times separately for each release location shows that anthrax, pneumonic plague, and tularaemia releases in Martin Place can be detected more quickly than the TTCD with few, if any, false alarms (see the supplementary materials). In contrast, at very low false alarm rates, Q fever releases in Martin Place are detected with worse lead times than Q fever releases in other locations. This is due to the substantially larger number of exposures in Martin Place, due to the population density, which in turn results in earlier TTCDs with less variance than for other release locations (see the supplementary materials).

Finally, we consider the trade-off between detection sensitivity and false alarm rates. As shown in Figure 7 (bottom panel), releases of anthrax, pneumonic plague, and tularaemia can be detected with very high sensitivity at low false alarm rates. In contrast, reliable detection of Q fever releases is only achieved at higher false alarm rates (e.g., 28 false alarms corresponds to one false alarm every 4 days). As highlighted in subsection 3.2, this is primarily due to the substantially smaller number of clinical events that occur in the Q fever releases. Consistent with detection timeliness, the highest sensitivity at low false alarm rates is achieved with higher population densities (see the supplementary materials).

## 4 Discussion

### 4.1 Primary findings

We have introduced an updated version of the EpiDefend outbreak detection system, and evaluated its performance in detecting simulated outbreaks of four different pathogens, embedded in retrospective real-world health surveillance data from the PEARL linked health database. High detection rates and low false alarm rates can be achieved for medium-sized releases of all pathogens except Q fever (releases of which cause far fewer clinical cases than for the other pathogens). For outbreaks of Q fever and tularaemia, EpiDefend can detect outbreaks at least as quickly as best-case estimates for the time to clinical detection (TTCD), and does so at extremely low false alarm rates. For inhalational anthrax, EpiDefend can detect outbreaks as fast as the TTCD, but with slightly higher false alarm rates. For pneumonic plague, EpiDefend can only match the TTCD with substantially higher false alarm rates. Releases in locations with higher population densities yield greater numbers of cases, and are easier to detect rapidly and with low false alarm rates.

### 4.2 Related studies

A wide range of methods for detecting disease outbreaks from surveillance data have been proposed [28–30]. Common statistical methods often define an expected number of cases over a given time period, and use a statistical test to identify whether the observed number of cases is significantly higher than the expected value (i.e., the goal is to identify when the data appear to be inconsistent with “normal” expectations). These methods typically assume that the data follow a specific distribution family and/or use predefined case thresholds to trigger an alarm. Bayesian methods have been proposed that take a broadly similar approach to EpiDefend, in that they estimate the likelihood that the data contain evidence of an outbreak (i.e., detecting a specific kind of signal in the data).

Due to the limited number of real-world outbreaks for which data are available and broadly consistent with current surveillance systems, these methods are most often evaluated using synthetic data, as highlighted by recent reviews in the field [4, 5, 31]. This is done by either adding simulated cases to real-world data [30] (as we have done here) or, more commonly, by simulating both the baseline and outbreak data [29].

One of the most commonly identified challenges is that of accurately characterising the expected number of cases in the absence of an outbreak. For example, Nordin et al. reported that the “relationship between the days of the week and detection of events is complex” [32], and Hogan et al. reported that they conditioned “on month of year and day of week to account for seasonal and day-of-week effects in historical surveillance data” [33]. In the context of using rapid (imperfect) diagnostic tests to detect measles outbreaks, Arnold et al. found that “detection performance is highly dependent on the structure and magnitude of the background ‘noise’ infections” [34]. Our use of an ensemble of regression forecasts to characterise this background signal reflects the current state of epidemic forecasting, where statistical models are highly competitive with, or out-perform, mechanistic models, and ensembles consistently out-perform individual models [35–38]. Because EpiDefend performs joint estimation of the background and release signals, it can also account for biases in the background forecast without necessarily inferring that a release must have occurred.

An open challenge for the field is to detect outbreaks in a manner that is (a) timely; (b) highly sensitive; and (c) highly specific. Consistent with our results for inhalational anthrax and pneumonic plague, existing studies have found it extremely difficult to match the time to clinical detection without extremely high false-alarm rates [31, 33, 39]. For example, Hogan et al. reported mean detection times of 1.11 days after the first ED presentation at a false-alarm rate of one per month, and 1.40 days at a false-alarm rate of one per three years [33], while we assumed a clinical detection would occur 15–19 hours after the first ED presentation. Similarly, Buckeridge et al. reported that “syndromic surveillance could detect an inhalational anthrax outbreak before clinical case finding”, but only when syndromic surveillance had extremely high specificity (≥ 0.9), corresponding to 1 false alarm every 10 days [39].

One advantage of EpiDefend over many of these detection methods is that it considers sliding windows of all possible starting times and durations over the 14-day inference window. Accordingly, the estimated probability of a release can accumulate gradually over time until sufficient evidence is observed, as we have shown for Q fever and tularaemia outbreaks. While this is a quantitative improvement over many other detection methods, our findings remain qualitatively consistent with existing studies: smaller, slower outbreaks are harder to detect, and require higher false-alarm dates to do so. Human interpretation and judgement remain key components of any operational outbreak detection system, as clearly demonstrated by the Electronic Surveillance System for the Early Notification of Community-Based Epidemics (ESSENCE), which is possibly the most widely-used operational system in the world today [40]. It combines a variety of temporal and spatial detection methods with custom queries and user-defined alerts and, unlike individual outbreak detection methods, aims to expose anomalies to users and enable them to find and monitor outbreaks. A similar workflow is possible with EpiDefend, because it can identify the observations that have had the greatest influence on the estimated probability of an attack (regardless of whether an alarm is triggered) and this could be used to flag specific observations for end-users to investigate.

### 4.3 Strengths

Here we have demonstrated that EpiDefend can improve detection times for biological attacks, using a rich real-world linked data set (PEARL) that captures details of all plausible clinical presentations resulting from an attack.

For each of the four pathogens considered in this project, simulated attacks were constructed by generated individual exposures using a detailed physical plume model, and simulating population mobility using a representative urban mobility data set. This allowed us to generate spatio-temporal profiles of symptom onset and healthcare presentation events. We have aggregated these healthcare events for the detection algorithm presented here, but the underlying attack simulations have a high level of individual detail that can be used in further iterations of the EpiDefend algorithm. In the results presented here, we have used these individual symptom profiles to define the clinical time to detection, based on the time of the first fulminant presentation.

We have separated the inference window (past 14 days) from the release simulation period (e.g., the 21-day window for Q fever releases). This may be useful for other attack scenarios where the consequences will not necessarily become immediately apparent in the relevant data streams (e.g., extended periods of exposure to low pathogen concentrations, pathogens with long incubation periods and/or gradual onset of disease), and avoids the challenge of having to generate longer-term predictions of what the past data should look like in the absence of an attack.

### 4.4 Limitations

While we have made use of rich health data sets and a highly detailed plume simulation model, the simulated healthcare events produced for each attack have relied on assumptions regarding population mobility and healthcare-seeking behaviours, for which there are limited, if any, data to inform or validate our choices.

We have used a representative mobility data set, the ABS 2016 Journey to Work, but this only captures a subset of urban mobility. Mobility data captured from other sources (e.g., telecommunications and social media platforms) were not used, due to challenges regarding data sensitivity, availability, and cost, but also based on concerns about the biases inherent in these data (personal communications, Cameron Zachreson). As explained above, we aggregated the healthcare events into daily counts, but subsequent iterations of EpiDefend will presumably include spatial features.

We have necessarily made assumptions about the probability that an individual will seek healthcare, given their current symptoms, and whether they will present to an emergency department or a general practice clinic. For simplicity, we have assumed here that all individuals who seek healthcare will do so at emergency departments, rather than making use of alternatives such as general practice (GP) clinics, home visit services, or telehealth platforms. While this is an overly optimistic assumption, in the absence of data to inform a precise value we argue that qualitatively this is a reasonable assumption for people who develop severe disease, particularly given the long delays in obtaining appointments in Australian public general practice clinics. For example, an Australian household survey in 2023-24 reported that 28% of people reported waiting longer than they felt acceptable for a GP appointment, 46% of people seeking urgent medical care waited ≥24 hours for a GP appointment, and 29% of people delayed or did not use GPs when they believed that medical care was needed [41].

We have also assumed that most, but not all, individuals who experience fulminant disease will arrive at an emergency department within 12 hours. This is possibly one of the most difficult assumptions to inform or validate, but EpiDefend’s detection sensitivity and timeliness should not be particularly sensitive to this assumption, since the delay between onset of fulminant disease and arriving at an emergency department has an identical impact on case visibility for EpiDefend and on the time to clinical detection benchmark (see Figure 3).

Recall that the time to clinical detection is defined relative to the first severe presentation, and occurs after a lag that characterises assessment and diagnosis delays. As we mentioned in subsection 2.7, in the event of a sufficiently large spike in unexplained fevers or respiratory presentations, metagenomics may be used to identify the responsible pathogen(s) [42, 43]. However, this is a complex decision process and we do not know what level of unknown fevers would trigger this response in a given hospital. Accordingly, we have not evaluated EpiDefend against detection using metagenomic techniques.

Detecting small attacks is fundamentally more difficult than detecting larger attacks, due to the decreased signal-to-noise ratio, and our results suggest that attack detection should be divided into two separate inference problems:

1. Whether there is a small attack, on the understanding that the estimated probability of an attack will rarely reach very high levels, and there will always be a substantial trade-off between early warning and false alarms;
2. Whether there is a substantial attack, for which it is reasonable to expect the estimated probability of an attack will become very high before a potential clinical detection, without incurring a high false-alarm rate.

Our results give a coarse indication of how small an attack we can reliably detect in a timely manner without incurring a high false-alarm rate, but a precise threshold requires further analysis.

### 4.5 Methodological extensions to EpiDefend

With the approach used in this project, we can divide the population into multiple hierarchies (e.g., by age, gender, location of residence) and use reconciled time-series forecasts to characterise the baseline healthcare-seeking propensity for each population group in a way that directly accounts for temporal effects such as seasonality, day-of-week effects, and gradual changes in case ascertainment. In particular, these population groupings need not be mutually exclusive — an individual may belong to multiple groupings, and contribute to the likelihood calculations for multiple data streams. These population groupings and the grouping-specific forecasts subsume the input nodes of the individual Bayesian network used in the original EpiDefend algorithm [10]. The background forecasting methods presented in this report provide a principled way to derive such forecasts for any target population that are well-characterised by appropriate surveillance data, and inherently capture temporal effects such as seasonality and trends in healthcare-seeking behaviours.

One challenge in making use of the richness in the linked PEARL data sets is deciding whether and how to incorporate specific details of individual healthcare events into the attack model likelihood functions. Examples of such refinements include: considering ambulance arrivals and admissions directly to critical care as separate signals; incorporating spatial structure into the observation models (e.g., looking for clusters of possibly-related cases); and developing methods to dynamically adapt the detection threshold in response to changes in the background health context that may not be easily incorporated into the background forecasts (e.g., accounting for rapid changes in the local context, such as the substantial number of human metapneumovirus cases reported in NSW in 2024 [44]).

Also worth noting is that this iteration of EpiDefend allows us to investigate the backcast observations likelihoods ℒ(**y_1_**_∶_**_t_**|**x_t_**) (see subsection 2.10) and identify the observations that had the greatest influence on the estimated probability of attack. While we have not made direct use of these insights in the analyses presented here, they should provide assistance and guidance when developing future iterations of EpiDefend. These insights are also extremely relevant to real-world deployment of EpiDefend, where they could help to identify cases linked to an attack, regardless of whether EpiDefend was able to detect the attack before clinical confirmation.

## 5 Conclusions

We evaluated the early-warning performance of EpiDefend using a real-world health data set that characterises healthcare demand in urban Australia. The results demonstrate that EpiDefend can reliably detect simulated attacks with delays comparable to expected clinical detection times, with extremely low false alarm rates. The implementation is computationally efficient and can be applied to health surveillance data in real-time; using only a single CPU core on the SURE platform, it consistently took 15–25 seconds (depending on the pathogen) to estimate the daily probability of an attack over each 14-day window. Furthermore, the methods are highly generalisable and could easily be adapted to other contexts, such as public health settings in other countries, and biological attack scenarios of relevance to national security.

From an operational perspective, the primary limitations of the EpiDefend algorithm and its performance are the assumptions concerning healthcare-seeking behaviours and mode of ED arrival, and it is unclear how these assumptions might possibly be informed or validated by existing real-world data. A truly robust evaluation of these assumptions would require a real-world release consistent with one of the attack models to occur, and for outbreak investigation and contact tracing efforts to characterise the disease progression and healthcare-seeking behaviours of all exposed individuals (noting that any such data collected during the COVID-19 pandemic would be inherently biased due to public awareness of COVID-19 prior to the onset of local transmission). Past outbreaks of pathogens such as *Legionella* (Legionnaires’ disease) [45] may also serve as real-world examples that EpiDefend should aim to detect with reasonable sensitivity, specificity, and timeliness. In addition to such retrospective analyses, real-world evaluation of EpiDefend’s performance could involve deploying it in an operational manner, parallel to an existing electronic disease syndromic surveillance system.

## Data Availability

The PEARL data are hosted on the Sax Institute's Secure Unified Research Environment (SURE). Approval to access these data must be obtained from data custodians at the New South Wales Ministry of Health, and applicants must have ethics approval.

## 6 Availability of materials

- The code for generating the background PEARL forecasts and the EpiDefend inference algorithm, and the simulated attack input data files, are available in a **currently private** git repository (https://gitlab.unimelb.edu.au/rgmoss/dst-epidefend-early-warning).
- The plume simulation data files (33.6 GB) are hosted on the University of Melbourne’s MediaFlux research data management platform.
- The PEARL data are hosted on the Sax Institute’s Secure Unified Research Environment (SURE). Approval to access these data must be obtained from data custodians at the New South Wales Ministry of Health, and applicants must have ethics approval.

## 7 Acknowledgements

RM was supported by DMTC Project 14.154 (EpiDefend: Disease Early Warning Tool). JMM is supported by an Australian Research Council (ARC) Laureate Fellowship (FL240100126). DJM is supported by an Australian National Health and Medical Research Council (NHMRC) Investigator Grant (APP1194109).

The contents of the published material are solely the responsibility of the Administering Institution, a Participating Institution or individual authors and do not reflect the views of the NHMRC. We thank the NSW Ministry of Health and The Centre for Health Record Linkage (CHeReL) for providing access to the data and syndrome definitions used in emergency department surveillance.

We thank Emma Prato and Maryanne Spiers at DMTC for overseeing the project, Jamil Ahmed and the staff at the Sax Institute for their support in using the SURE platform, and Dr Cameron Zachreson at the University of Melbourne for recommending the ABS JTW data over other mobility data sources. Finally, we thank the clinicians and microbiologists whose expert advice informed the time to clinical detection estimates used in this project, including: Danny Liew and Vincent Sinickas (Royal Melbourne Hospital), Allen Cheng (Alfred Health), Matthias Dorsch and Craig Brinkworth (DST).

## A Supplementary Materials: Methods

### A.1 Characterise normal health signals

The aim of EpiDefend is to estimate the probability that among the cases reported in health data over the past 14 days, some of these cases are due to a biological attack. We assumed that health data before this 14-day period do not include any cases arising from a biological attack, as it is expected that an attack from more than 14 days in the past would have been detected by pathological testing. Accordingly, we treated the health data for the past 14 days as suspect, and used an ensemble of regression models to forecast each of the six data streams (defined in the previous section) based on the health data up to 14 days ago. We generated these forecasts for every 14-day window in the study period.

Forecasts for the age-aggregated syndromic EDDC presentations were generated using an ensemble of two different models, and seven model variants in total:

- A seasonal naive model: an ARIMA model with a white noise non-seasonal component, and a random walk seasonal component. This is the seasonal version of the standard naive forecast (random walk).
- An exponential smoothing model with trend and season components (ETS), using six different reconciliation methods to ensure consistency between the individual forecasts for each age group and the all-age forecast:

**–** Bottom-up: all-ages counts are the sum of the individual age-group counts;
**–** Minimum trace [46]: minimise total forecast variance using different estimates of forecast error variance: ordinary least squares (OLS), weighted least squares using variance scaling (WLS var), weighted least squares using structural scaling (WLS struct), sample covariance (MinT cov), and sample covariance shrinkage estimator (MinT shrink).

Forecasts for the critical admissions were generated using a single ETS model, which consistently out-performed the seasonal naive model. Because these data are a single time-series, no reconciliation was required. We applied a power transformation to ensure that the seasonal variation remained consistent across the entire time-series, using the Guerrero feature [47] to choose an appropriate value for the power parameter.

For each 14-day window, each of the above models were trained on the aggregated time-series data up to the start of inference/detection window, and were then combined into an equal-weights ensemble.

### A.2 Chosen pathogens

Because the PEARL data captures respiratory infections, and viral pathogens of concern can be rapidly identified using RT-PCR assays, we chose four bacterial pathogens that are plausible bioterrorism candidates and which result in symptoms consistent with common respiratory infections: anthrax, pneumonic plague, Q fever, and tularaemia.

#### A.2.1 Anthrax

We characterised the stages of an anthrax infection with log-normal distributions, whose parameters are defined as per previous iterations of the EpiDefend algorithm [10]:

**Anthrax incubation period** Lognormal(𝜇 = log(5), *σ* = 0.4)

**Anthrax prodromal period** Lognormal(𝜇 = log(2.3), *σ* = 0.3)

#### A.2.2 Pneumonic plague

We characterised the incubation stage of a pnuemonic plague infection with a log-normal distribution, whose parameters are defined as per previous iterations of the EpiDefend algorithm [10]:

**Pnuemonic plague incubation period** Lognormal(𝜇 = log(4), *σ* = 0.4)

#### A.2.3 Q fever

The incubation period for acute Q fever was informed by a recent systematic review [48], which reported a median incubation period of 18 days, with 95% of cases expected to occur between 7 and 32 days after exposure. We fitted a log-normal distribution to this median and 95% interval:

**Q fever incubation period** Lognormal(𝜇 = log(18), *σ* = 0.343)

Another review article reported that symptom onset is usually abrupt, “with severe fever, fatigue, chills, and headaches”, and that the clinical presentations most often present with febrile illness and/or pneumonia [49]. Due to the sudden onset of severe symptoms, we assumed that infected persons would rapidly seek healthcare in response to symptom onset, and did not model subsequent disease progression.

#### A.2.4 Tularaemia

The WHO Guidelines on Tularaemia [50] report a mean incubation period of 3–5 days, with a range of 1–21 days, and that “onset of disease is abrupt, including rapid development of fever with chills, fatigue, general body aches and headache”. We assumed that the incubation period could be characterised by a log-normal distribution with a median of 4 days, and that for 95% of cases the incubation period is between 1 and 21 days.

**Tularaemia incubation period** Lognormal(𝜇 = log(4), *σ* = 0.76)

The guidelines also report that in type A tularaemia (the strain that is most likely to be used as a biological weapon), “pneumonia is a fulminant condition” and that the condition “is extremely severe”. Due to the sudden onset of severe symptoms, we assumed that infected persons will rapidly seek healthcare in response to symptom onset, and did not model subsequent disease progression.

### A.3 Spatial population structure

We used Australian Bureau of Statistics (ABS) Journey to Work (JTW) data to characterise population mobility in metropolitan Sydney. An individual’s risk of exposure to a pathogen release, and their subsequent location when symptom progression causes them to seek healthcare, are informed by these mobility data, reported per ABS Statistical Areas Level 1 (SA1) and Level 2 (SA2) for metropolitan Sydney [51], and ABS estimated resident populations for each SA2 [52]. Given an individual’s location when they seek healthcare, we then select the nearest facility from a data set of Australian hospital locations [22]. Note that the JTW data were retrieved from the ABS using their TableBuilder tool [53], from which we obtained counts of employed persons aggregated by place of usual residence (PUR) and place of work (POWP). These data were most recently collected in the 2016 Census, and for consistency we have used 2016 data sets for statistical area definitions, boundaries, and estimated resident populations.

### A.4 Time to clinical detection

For each pathogen, we defined delay distributions to estimate the time at which a public health clinician would be likely to identify the release pathogen in a clinical case. These delay distributions are the benchmarks against which the detection performance of the EpiDefend algorithm was compared and evaluated. Pathogen-specific models for the time to clinical detection (TTCD) were informed by information collecting from a variety of sources:

1. Meetings held with clinicians and pathologists at the Royal Melbourne Hospital and The Alfred Hospital;
2. Surveillance and public health laboratory guidelines published by the Australian Commonwealth Department of Health and Departments/Ministries of Health in individual Australian states; and
3. Relevant peer-reviewed articles published in medical and scientific journals.

Each pathogen-specific model shares the same sequence of events that are necessary for a clinical detection to occur, and the differences between the pathogen-specific models are captured in the delay distributions for disease progression and laboratory testing (see Figure A1).

**Figure A1:**
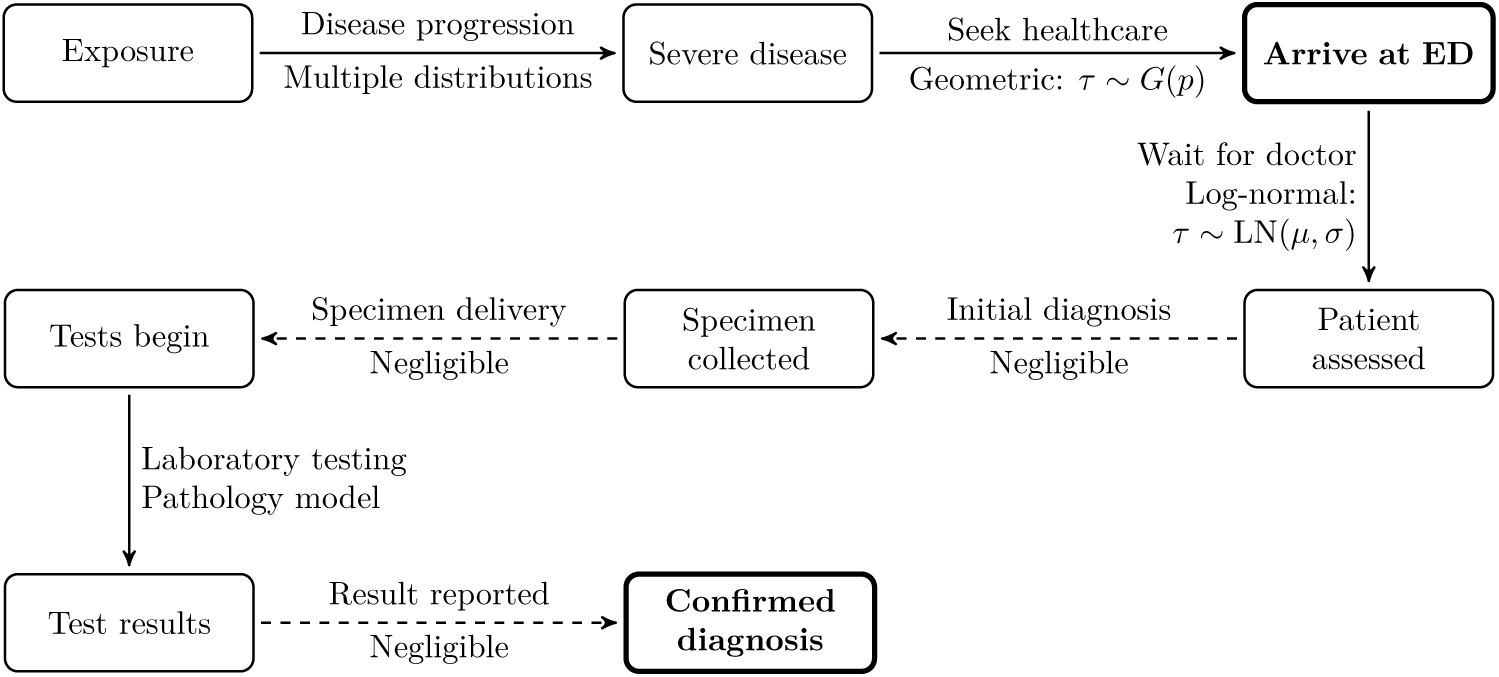
A diagram of the sequence of events that begins with an initial exposure and culminates in a clinical detection of the release. The time to clinical detection is the delay between arriving at the ED (bold node) and a confirmed diagnosis (bold node).

The delay between arriving at an Emergency Department and being assessed by a clinician was informed by a 2009 study of 501 pneumonia patients in an Australian emergency department, which reported the delay between the time of presentation to the hospital triage desk, and the documented time of first antibiotic administration in the medication chart [24]. Delays were reported for each triage severity class, and we assumed that patients with severe respiratory symptoms would be triaged as either severity class 4 or severity class 5:

- Severity class 4: median 2 hours, IQR of 1 to 4.2;
- Severity class 5: median 3.1 hours, IQR of 1.8 to 4.5.

Accordingly, we assume a median of 2.5 hours and IQR of 1.4 to 4.3. The study also reported CDF point estimates for patients who had not previously received antibiotics:

- 175 (35%) received antibiotics within 2 hours of presentation;
- 346 (69%) received antibiotics within 4 hours; and
- 454 (91%) received antibiotics within 8 hours.

We obtained a good fit to these data points with a log-normal distribution:

**Delay** Lognormal(𝜇 = log(2.5), *σ* = 0.85)

**Figure A2:**
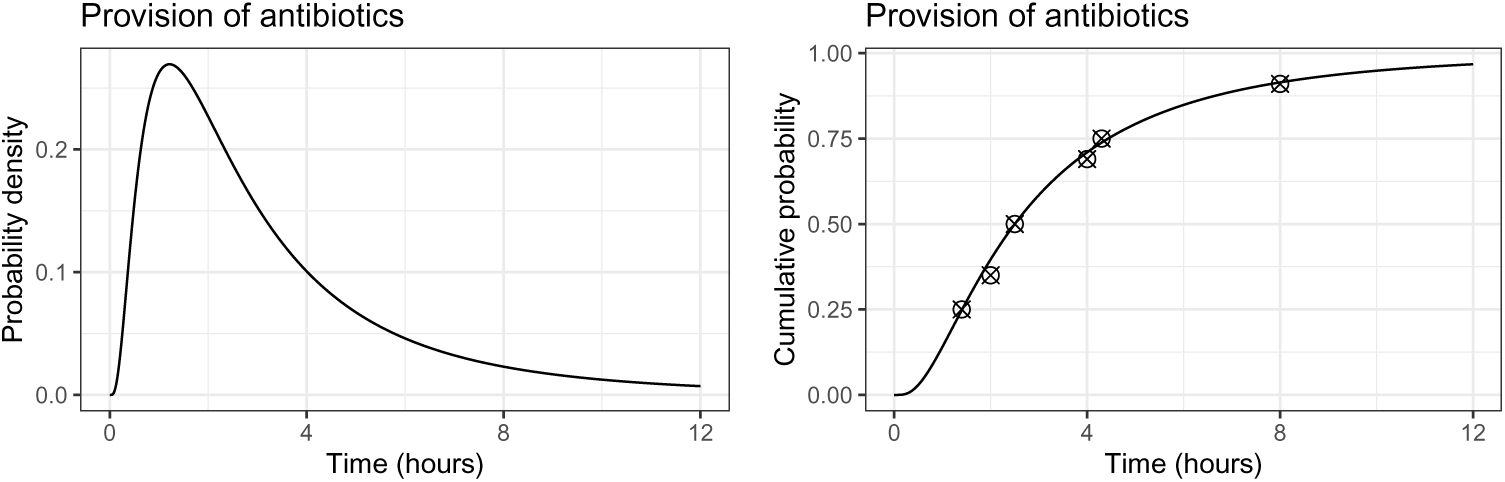
The delay in provision of antibiotics to pneumonia patients„ shown as a probability density (left panel) and a cumulative distribution (right panel). The cumulative distribution is compared to several reported observations (crossed circles).

The estimated laboratory testing delays were:

**Anthrax** 15–19 hours, based on a culture time of 12 hours, 20 minutes to obtain results via MALDI-TOF, and 3–5 hours to redo the test and further consultation.

**Pneumonic plague** 15–19 hours, based on a culture time of 12 hours, 20 minutes to obtain results via MALDI-TOF, and 3–5 hours to redo the test and further consultation.

**Q fever** 24–48 hours by RT-PCR, but this is unlikely in an urban area; serology would be the most likely diagnosis method, with a delay of up to 10 days after symptom onset

**Tularaemia** ≥7 days, due at least in part to limited awareness. There is no serology testing in Australia, RT-PCR testing is limited, and culture is difficult and not routine; routine blood cultures will not return a positive result.

In an urban mass-exposure event, neither Q fever or Tularaemia would be detected in a timely manner by pathology laboratories. Hypothetically, if a hospital experienced a sufficiently-large spike in unexplained fevers, they would rely on genetic sequencing to identify the responsible pathogen(s). However, we do not know what level unknown fevers would trigger this response.

To avoid evaluating EpiDefend’s performance against such benchmarks, where success is all but guaranteed, we have instead assumed best-case diagnosis delays of 48 hours for Q fever (assuming positive RT-PCR results within several days of symptom onset) and 72 hour for tularaemia (assuming that the pathology laboratory uses MALDI-TOF mass spectrometry, and that the MALDI-TOF library contains a Francisella entity).

### A.5 Disease models

To simulate disease progression and hypothetical healthcare events in the EpiDefend algorithm, we used a stochastic SEIR model with flexible numbers of *E* and *I* compartments (allowing us to fit the natural disease parameters of each pathogen) with mean transition rates defined by the ODE system:

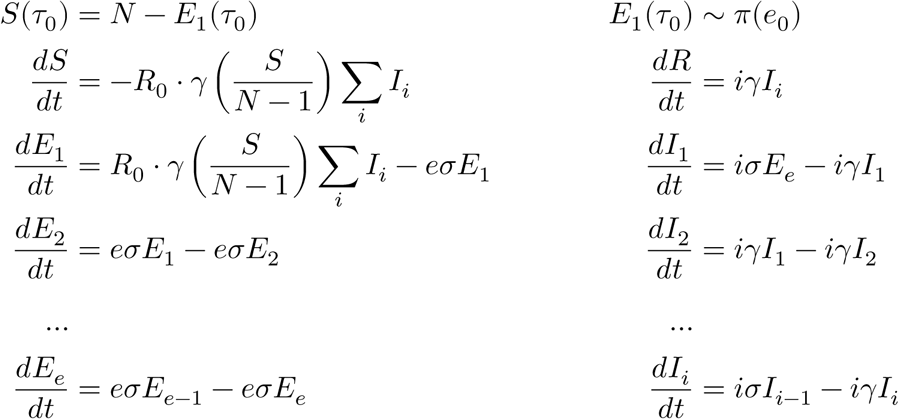

Of the four pathogens, only pneumonic plague allows for human-to-human transmission; for the other pathogens, we assumed *R*_0_ = 0. The E and I compartments were used to represent the incubation and prodromal stages of disease, and the R compartment represented the end of the prodromal stage (i.e., either the onset of fulminant disease, or recovery from infection for pathogens where this is a possible outcome). For the pathogens that do not include a prodromal stage (pneumonic plague, Q fever, and Tularaemia), we used a single I compartment to represent the final pre-fulminant stage. To determine the number of E and I compartments for each pathogen, we fit Gamma distributions with integer shape parameters to each of the disease distributions presented in subsection A.2. The number of compartments, and corresponding rate parameters, are listed in Table A1, and the prior distributions for the number of initial exposures are listed in Table A2.

**Table A1:** The SEIR model structure for each pathogen: the number of E and I compartments, the rate parameters for these compartments, and the nth I compartment at which prodromal cases may present (only for pathogens with a prodromal stage).

| Pathogen | # E | $\sigma$ | # I | $\gamma$ | # I prodromal |
| --- | --- | --- | --- | --- | --- |
| Anthrax | 6 | 0.903 | 11 | 0.219 | 6 |
| Plague | 5 | 1.391 | 1 | 1.391 | — |
| Q fever | 7 | 2.39 | 1 | 2.39 | — |
| Tularaemia | 1 | 2.68 | 1 | 2.68 | — |

**Table A2:** The prior distributions 𝜋(*e*_0_) for initial exposures, for each pathogen and each release size. Differences in prior distributions between the pathogens reflect the range of simulated cases arising from each simulated release.

| Pathogen | Small | Medium | Large | Huge |
| --- | --- | --- | --- | --- |
| Anthrax | $\mathcal{U}(50, 200)$ | $\mathcal{U}(200, 1000)$ | $\mathcal{U}(1000, 1800)$ | $\mathcal{U}(1800, 2500)$ |
| Plague | $\mathcal{U}(50, 200)$ | $\mathcal{U}(200, 1000)$ | $\mathcal{U}(1000, 1800)$ | $\mathcal{U}(1800, 2500)$ |
| Q fever | $\mathcal{U}(5, 50)$ | $\mathcal{U}(50, 500)$ | $\mathcal{U}(500, 1000)$ | $\mathcal{U}(1000, 2500)$ |
| Tularaemia | $\mathcal{U}(10, 50)$ | $\mathcal{U}(100, 1000)$ | $\mathcal{U}(1000, 1800)$ | $\mathcal{U}(1800, 2500)$ |

The particle state vector comprised the time of the release *τ*_0_, the number of initial exposures *e*_0_, the SEIR model parameters and state variables, and a forecast trajectory index *f*:

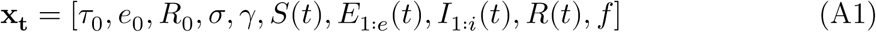

### A.6 Likelihood

Observable healthcare events were arise from both prodromal and fulminant presentations. For anthrax infections, we assumed that prodromal presentations occurred at the midpoint of the prodromal stage (i.e., when an individual enters compartment *I*_6_), and we set the prodromal visibility index 𝑣 = 6. For all other pathogens, there is no prodromal stage and we set 𝑣 = *i* + 1. With this index 𝑣 we defined the subset *V* of all particles **x_t_** for which there has been at least one infection that may have resulted in an observed case (A2), and sum the weights 𝑤_*i*_ of all particles in *V* to define the probability of a visible attack at time *t* (A3):

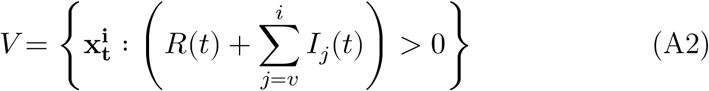

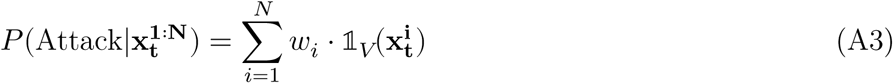

The expected value 𝐵_*f*_ for data stream *d* with forecast trajectories {*d*_1_, *d*_2_, …, *d*_𝑘_} at time *t*, in the absence of an attack, depends on each particle’s forecast trajectory index *f*:

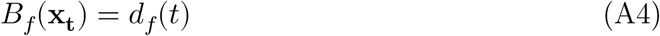

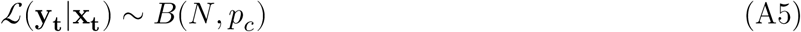

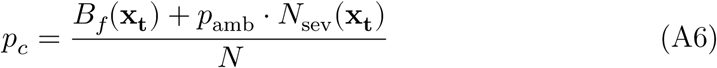

For age-specific and all-age respiratory EDDC presentations, the likelihood of observing **y_t_**cases was modelled as a negative binomial process with dispersion parameter 𝑘, where we expect a proportion *p*_mild_ of the *N*_mild_ prodromal cases to present, and assume a fraction *A* of exposures occur in this age group:

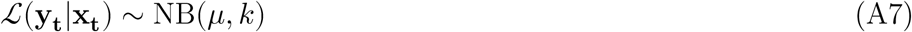

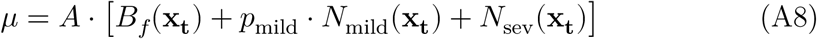

The number of prodromal cases over the past day is the number of individuals who entered the *I*_𝑣_ compartment, for pathogens that include a prodromal stage:

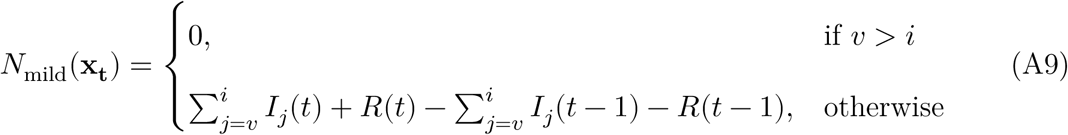

The number of severe presentations over the past day is a more complex quantity to calculate, due to the geometric delay distribution for the time between onset of fulminant disease (entering the *R* compartment) and arriving at a hospital. We consider individuals who entered *R* in each of the past 48 hours, and for each hourly window calculate the probability that these individuals would have arrived at a hospital in the past 24 hours:

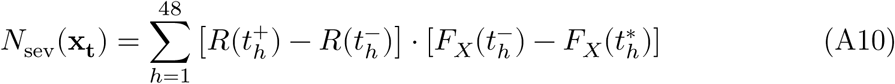

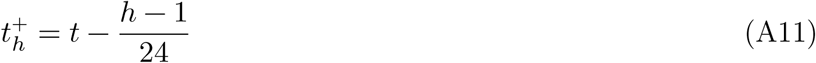

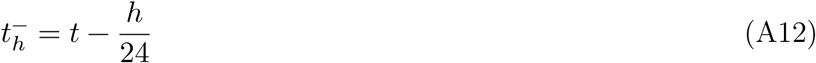

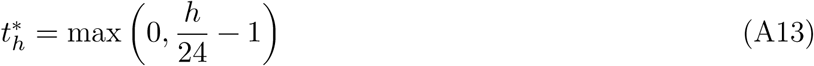

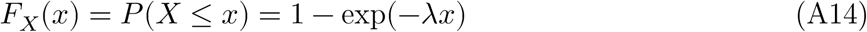

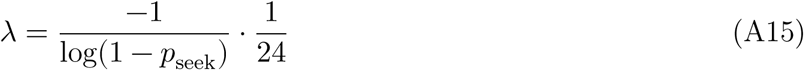

## B Supplementary Materials: Results

### B.1 Pathogen releases and healthcare events

Simulated healthcare events for small and medium releases of each pathogen are shown in Figure B1 and Figure B2, for each combination of release location and release hour (i.e., time of day). For each release, there is a simulated time-series of daily EDDC presentations with symptoms consistent with one of the PEARL respiratory syndromes, and a simulated time-series of daily EDDC ambulance arrivals who are directly admitted into critical care. A comparison of these simulated attack time-series demonstrates the substantially higher population density in the Sydney CBD (Martin Place) relative to the other release locations. A small release in Martin Place typically produces a similar number of cases to a medium release in Parramatta, and a greater number of cases than medium releases in the other locations.

**Figure B1:**
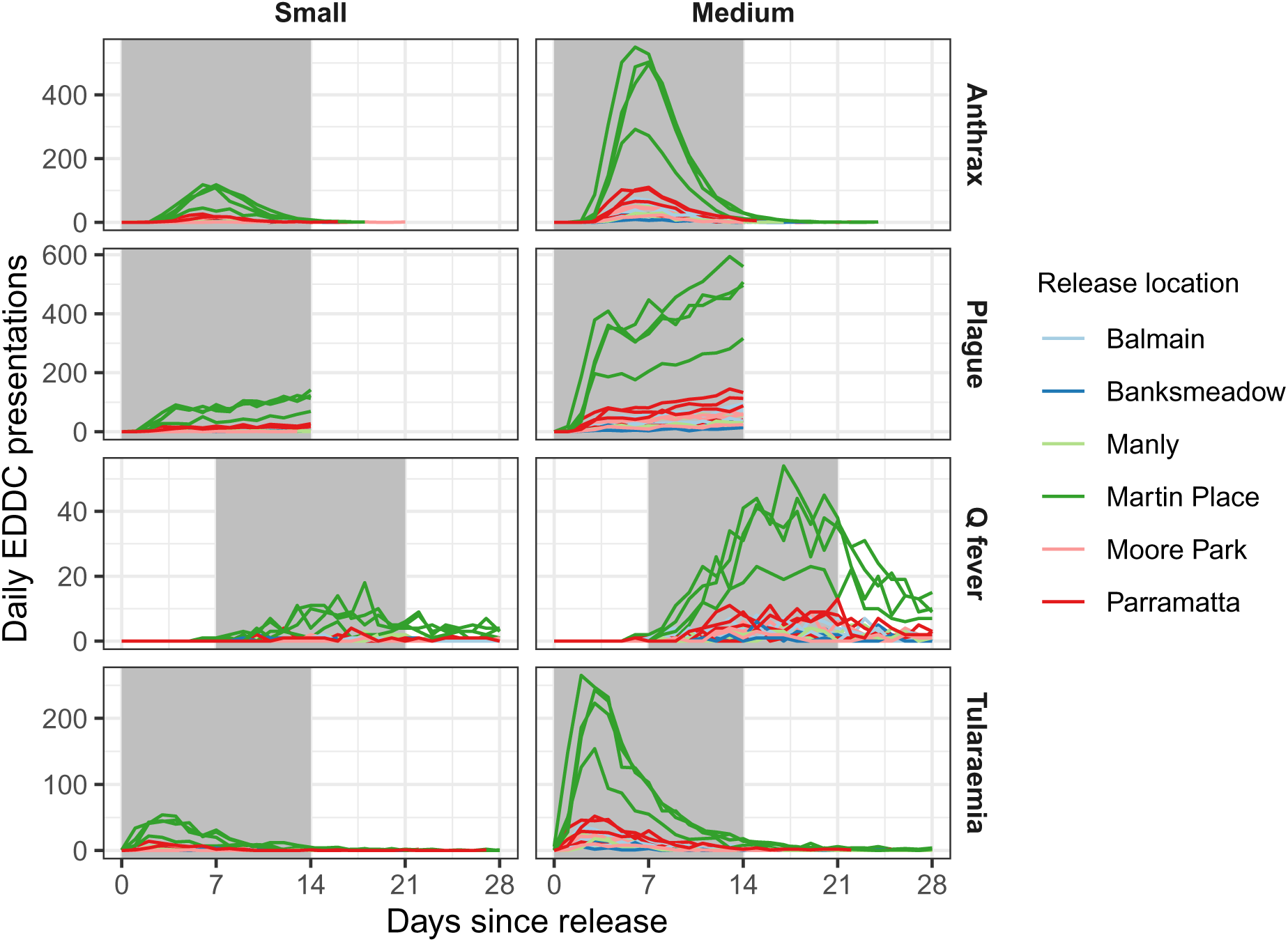
Simulated emergency department respiratory presentations for each small and medium pathogen release. Shaded grey regions indicate the time period over which simulated events are injected into the PEARL data.

**Figure B2:**
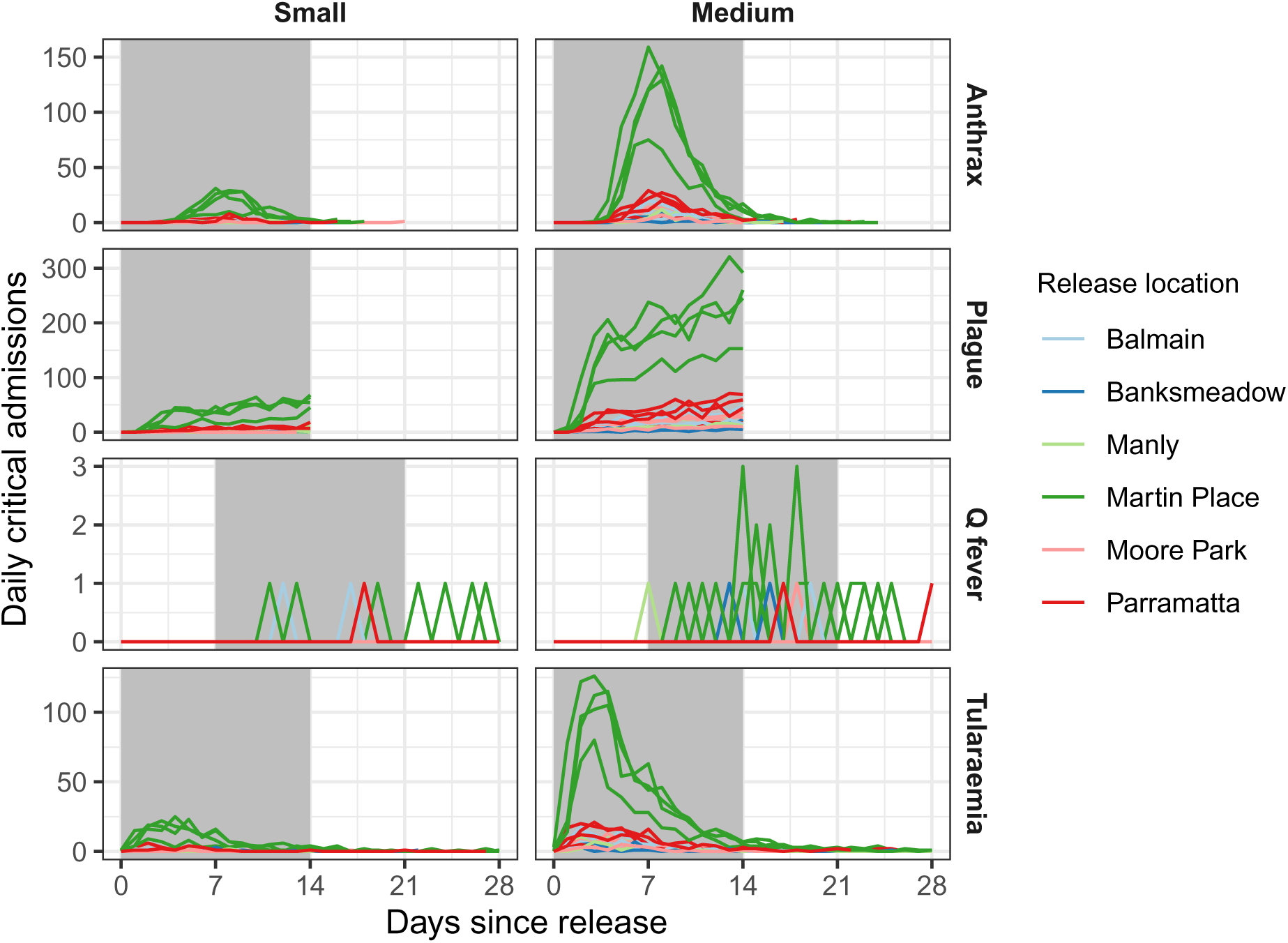
Simulated critical respiratory admissions for each small and medium pathogen release. Shaded grey regions indicate the time period over which simulated events are injected into the PEARL data.

For all pathogens except Q fever, small releases in Martin Place and medium releases in all locations produce enough critical admissions that this signal, in isolation, should be sufficient for EpiDefend to detect a release *at some point*. But this does not guarantee that EpiDefend will detect the release before a clinical diagnosis is made, because the time to clinical detection is defined relative to the first clinical presentation — which will occur before, or at the same time as, the first critical admissions caused by the release.

### B.2 Time to clinical detection

The simulated times to clinical detection for medium releases of each pathogen are shown in Figure B3. Recall that these are defined relative to the first presenting case. Releases in the Sydney CBD (Martin Place) exhibit the smallest variance in detection times, and have slightly lower mean times to detection than releases in the other locations. For medium releases, mean detection times are 5.6 days (anthrax), 2.9 days (pneumonic plague), 17.8 days (Q fever), and 4.6 days (tularaemia).

**Figure B3:**
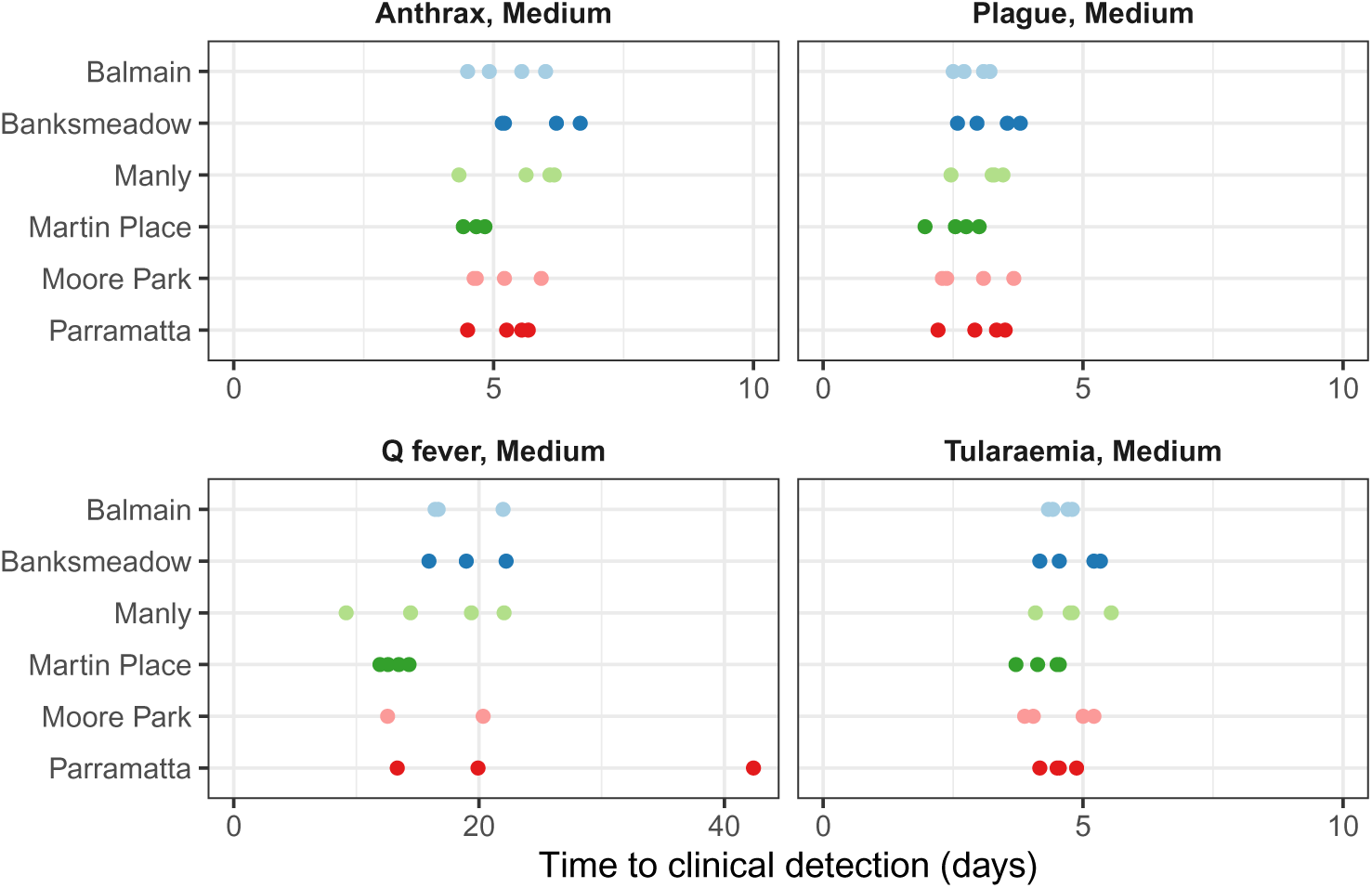
The simulated times from pathogen release to clinical detection, for medium releases of each pathogen, shown separately for each release location and release hour.

Note that several of the Q fever releases do not result in a clinical detection (1 release in each of Balmain, Banksmeadow, and Parramatta; 2 releases in Moore Park). This is because all of the simulated exposures yielded either no symptoms or only acute self-limited symptoms, and so a positive diagnosis would be extremely unlikely in an urban area. When calculating Epidefend’s lead time relative to clinical detection, we did not include these releases.

Small releases have the longest times to clinical detection, and detection times decrease as the release size increases. Mean detection times for small and huge releases are: 6.0 days and 4.3 days for anthrax, 3.6 and 2.4 days for pneumonic plague, 17.4 and 11.1 days for Q fever, and 5.1 and 4.1 days for tularaemia.

### B.3 PEARL data and background forecast skill

We used an ensemble of seven model variants to forecast the age-aggregated syndromic EDDC presentations. Example forecasts are shown against the ground truth PEARL data in Figure B4, for several 14-day forecast horizons in the 2018 winter. With respect to the 2018 winter, and to the winter periods of the 2016–2019 calendar years, forecasts were generally in good agreement with the PEARL data, but exhibited short periods of systematic bias. As shown in Figure B5, no model variant consistently out-performed the other model variants; the ETS model variants generally out-performed the seasonal naive model, but on occasion the ETS models performed noticeably worse than the seasonal naive model. For this reason, we included the seasonal naive model in the ensemble.

**Figure B4:**
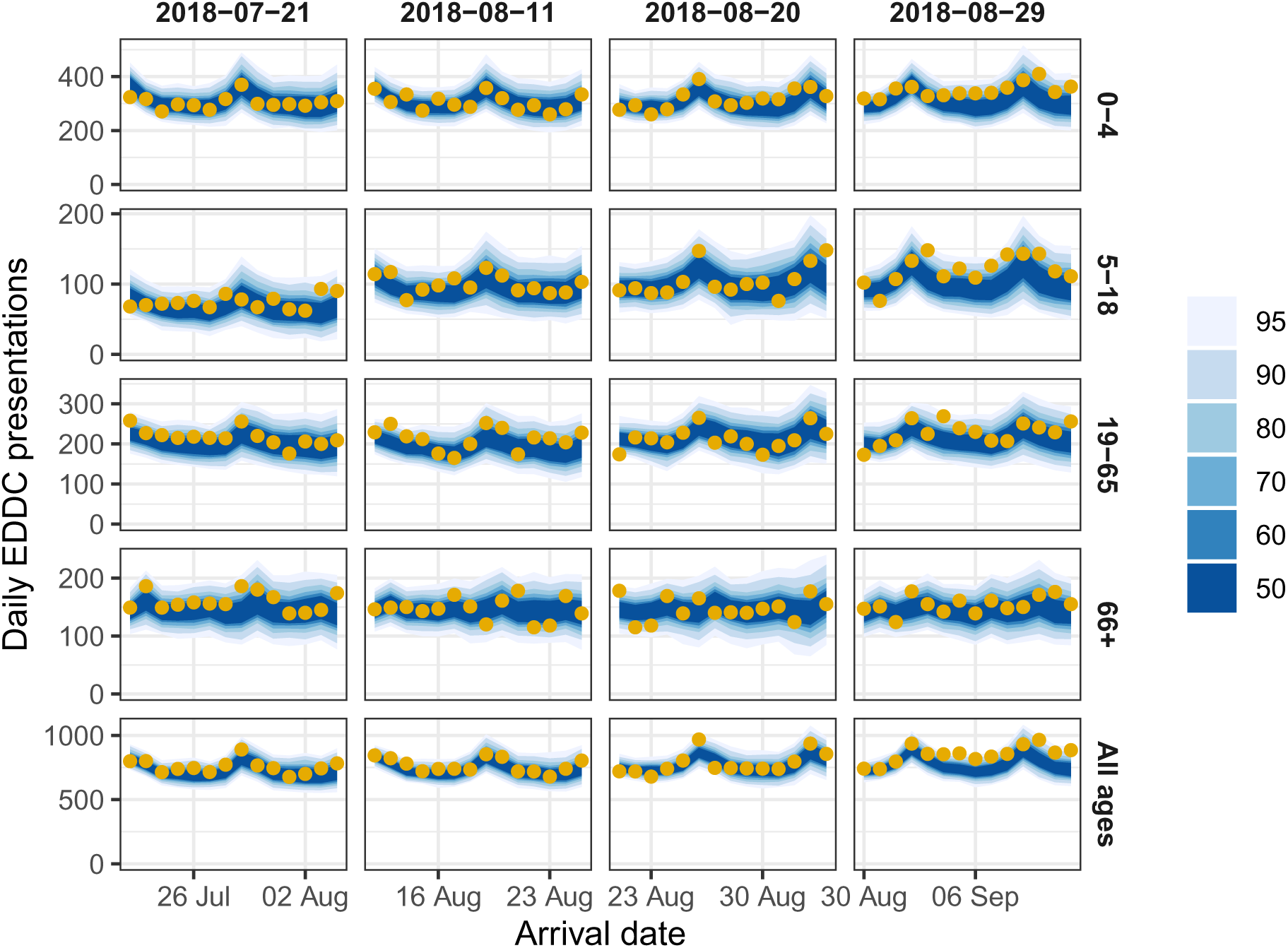
Daily counts of syndromic EDDC presentations (yellow points) shown for each age group and for all ages (rows), and 14-day ensemble forecast credible intervals for these data (blue regions). Data and forecasts are shown for four different 14-day windows (columns) in the 2018 winter.

**Figure B5:**
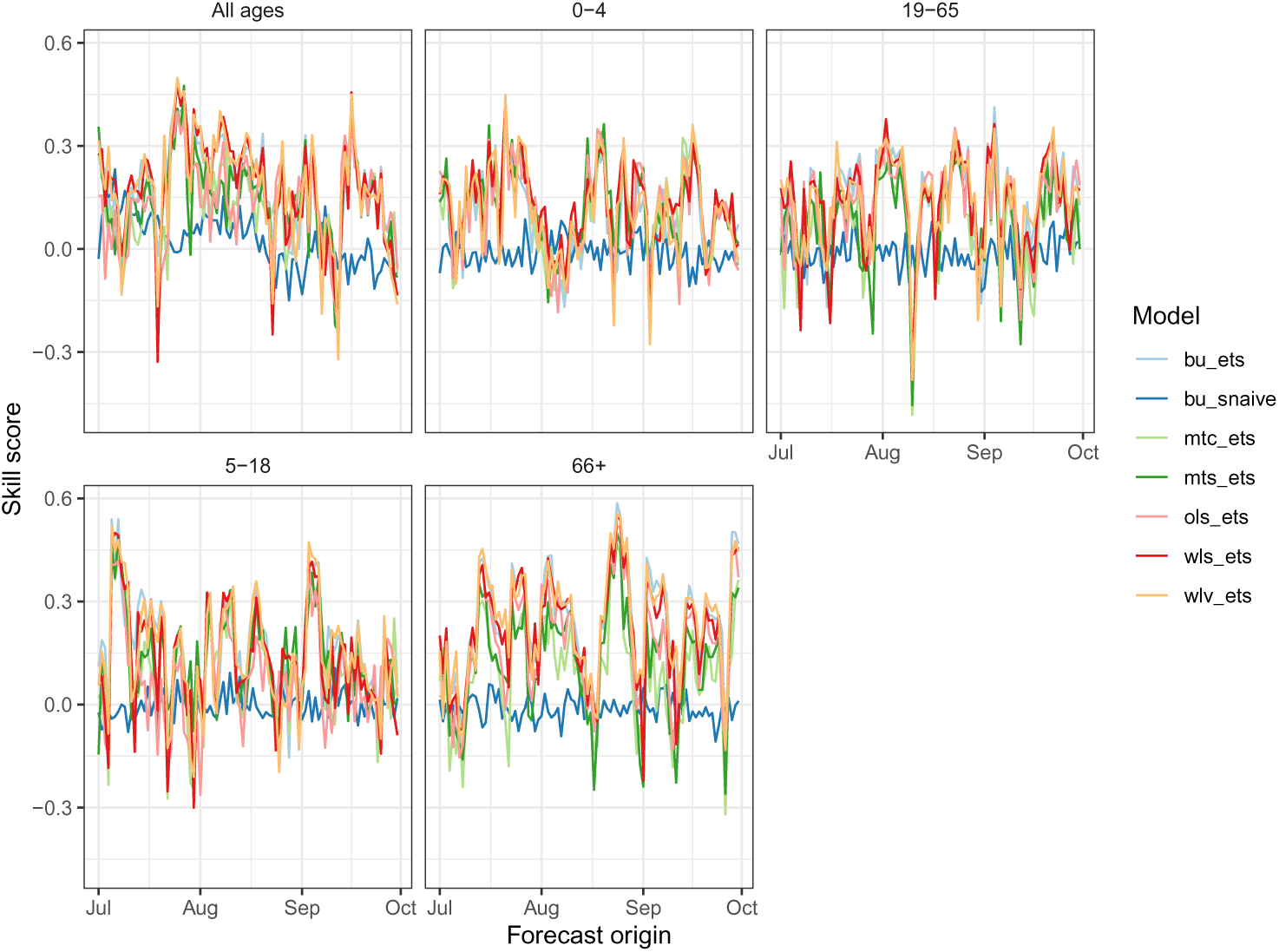
Skill scores for each model variant over the 2018 study period, shown separately for each age-group and for all ages.

The “Critical admissions” time-series (patients who arrive by ambulance and are admitted directly into critical care) was relatively stable over the study period, in comparison to the syndromic EDDC presentations. As shown in Figure B6, the ETS model consistently out-performed the seasonal naive model. Its skill score was almost always positive, and only ever dropped slightly below zero. For this reason, we only used the ETS model to forecast the severe event counts. The 14-day forecasts were consistently in good agreement with these data, as shown in Figure B7.

**Figure B6:**
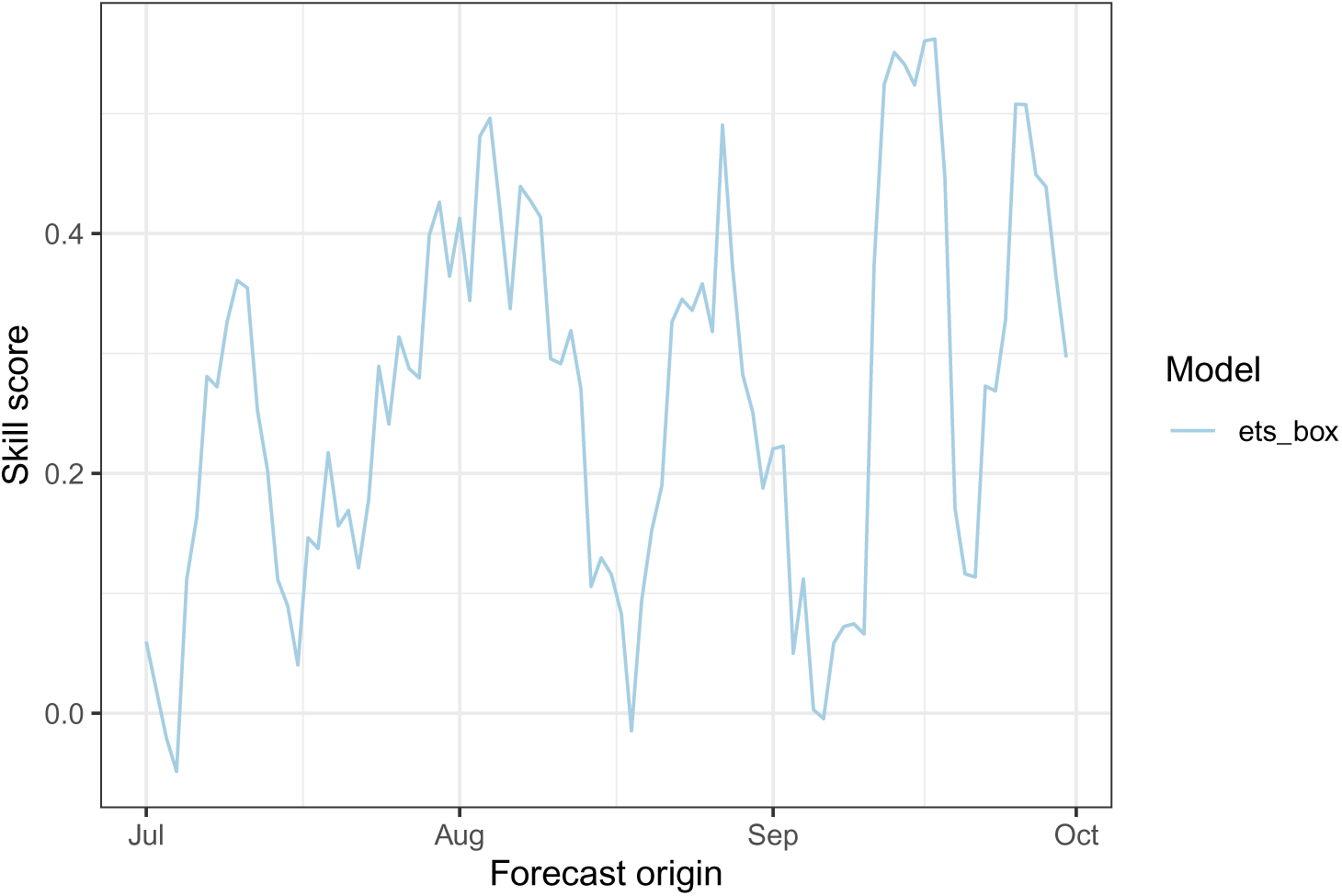
Skill score of the ambulance to critical care forecasts over the 2018 study period.

**Figure B7:**
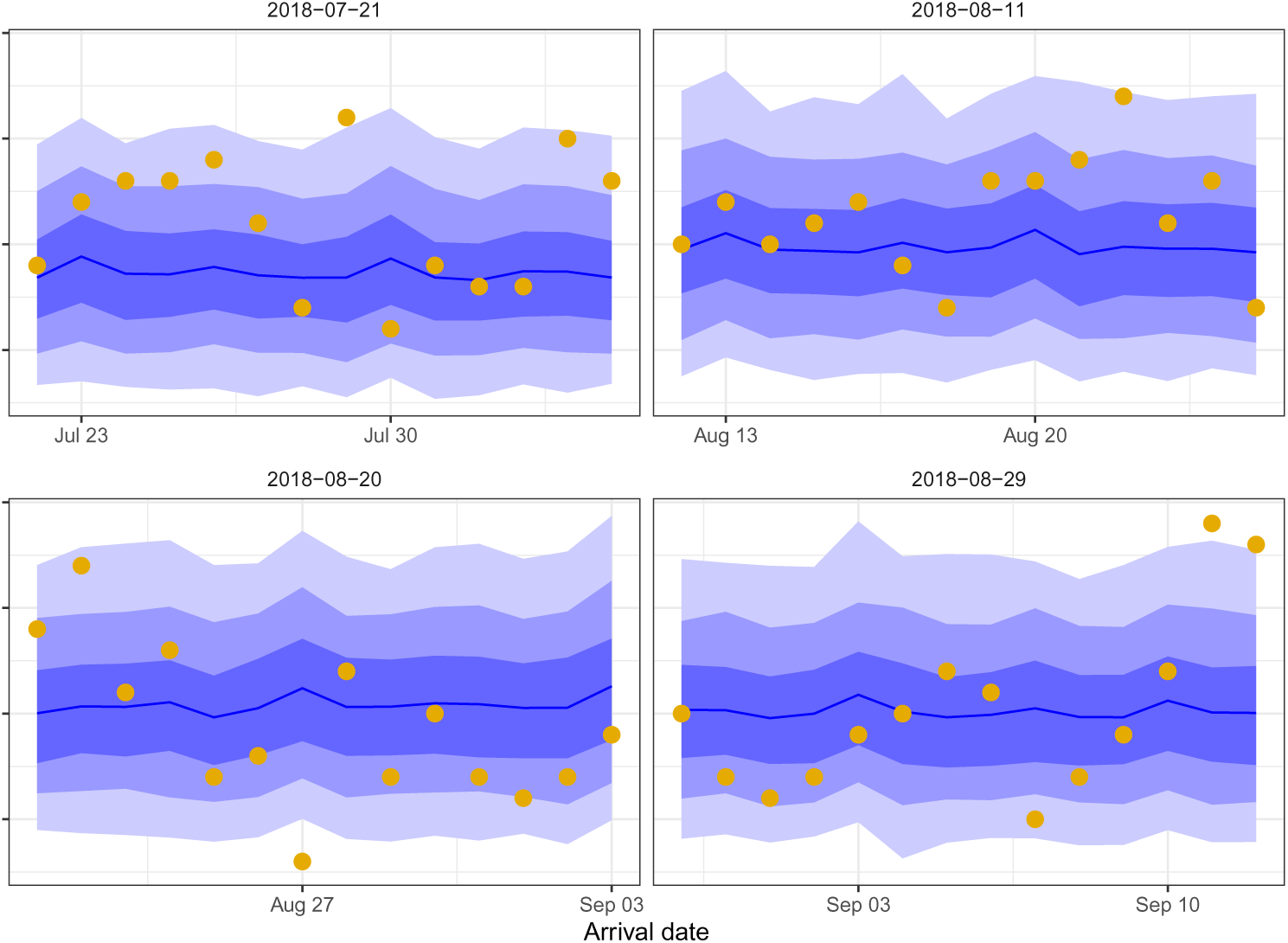
Daily counts of syndromic EDDC presentations that arrive via ambulance and are directly admitted to critical care (yellow points) and 14-day forecast credible intervals for these data (blue regions). Data and forecasts are shown for four different 14-day windows (columns) in the 2018 winter. Note that the y-axis scale is omitted, due to the small counts in the time-series.

### B.4 EpiDefend sensitivity and specificity

The detection performance for each release location is shown in Figure B8.

**Figure B8:**
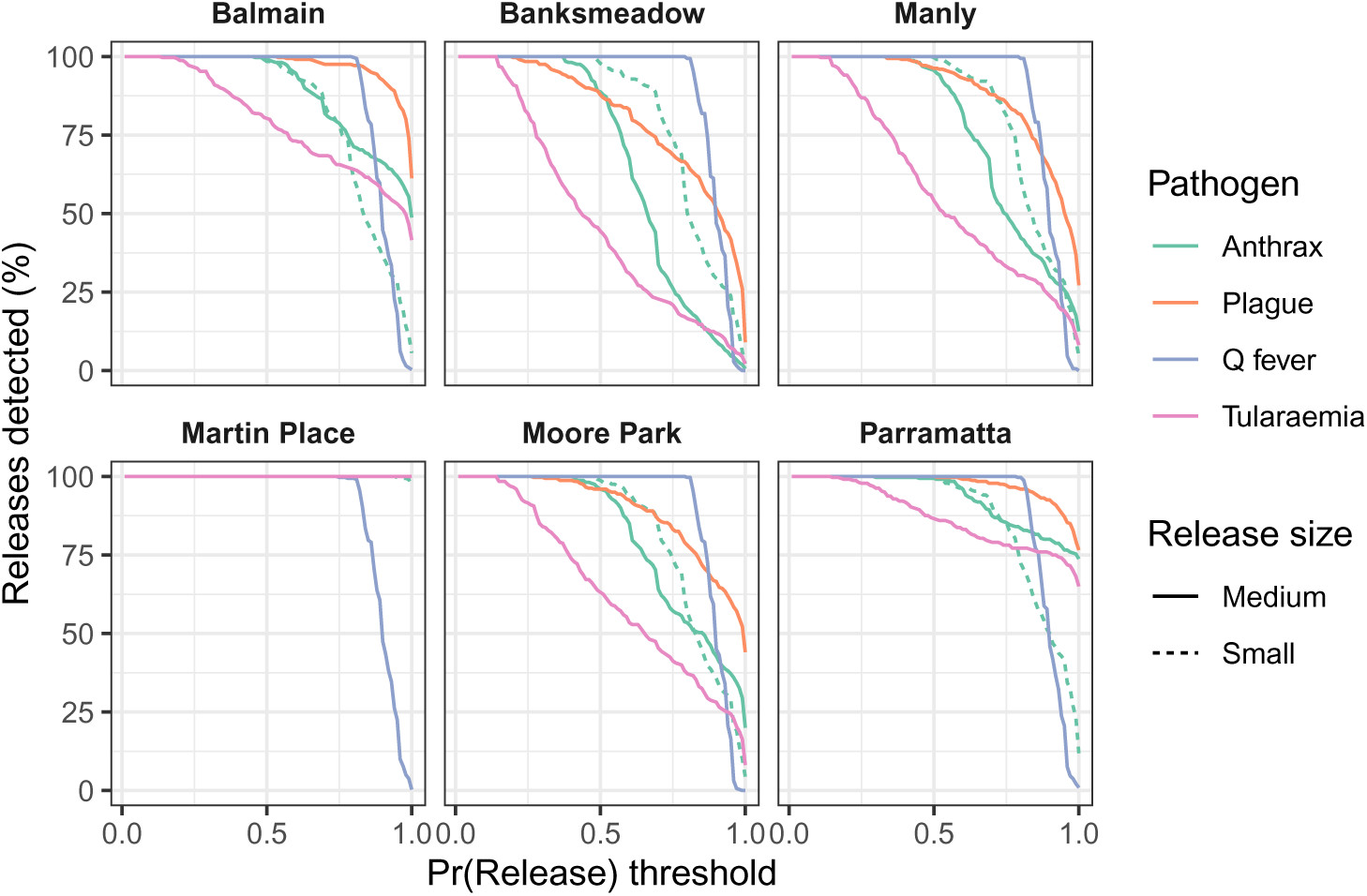
The percentage of simulated attacks that were detected, shown for each detection threshold, and reported separately for each release location.

### B.5 EpiDefend detection timeliness

We now demonstrate the trade-off between detection timeliness and false alarm rates with Activity Monitor Operator Characteristic (AMOC) curves. Figure B9 shows the relationship between the false alarm rate and the mean delay in detecting an attack (measured from the time of pathogen release) for each pathogen. The shortest detection delays are achieved with high false alarm rates (e.g., 57 false alarms corresponds to one false alarm every 2 days). Lower false alarm rates result in longer detection delays, and for zero false alarms the detection delays are approximately double the shortest detection delays. With the exception of Q fever, pathogen releases in the Sydney CBD (Martin Place) are consistently detected earlier than releases in the other locations (Figure B10).

**Figure B9:**
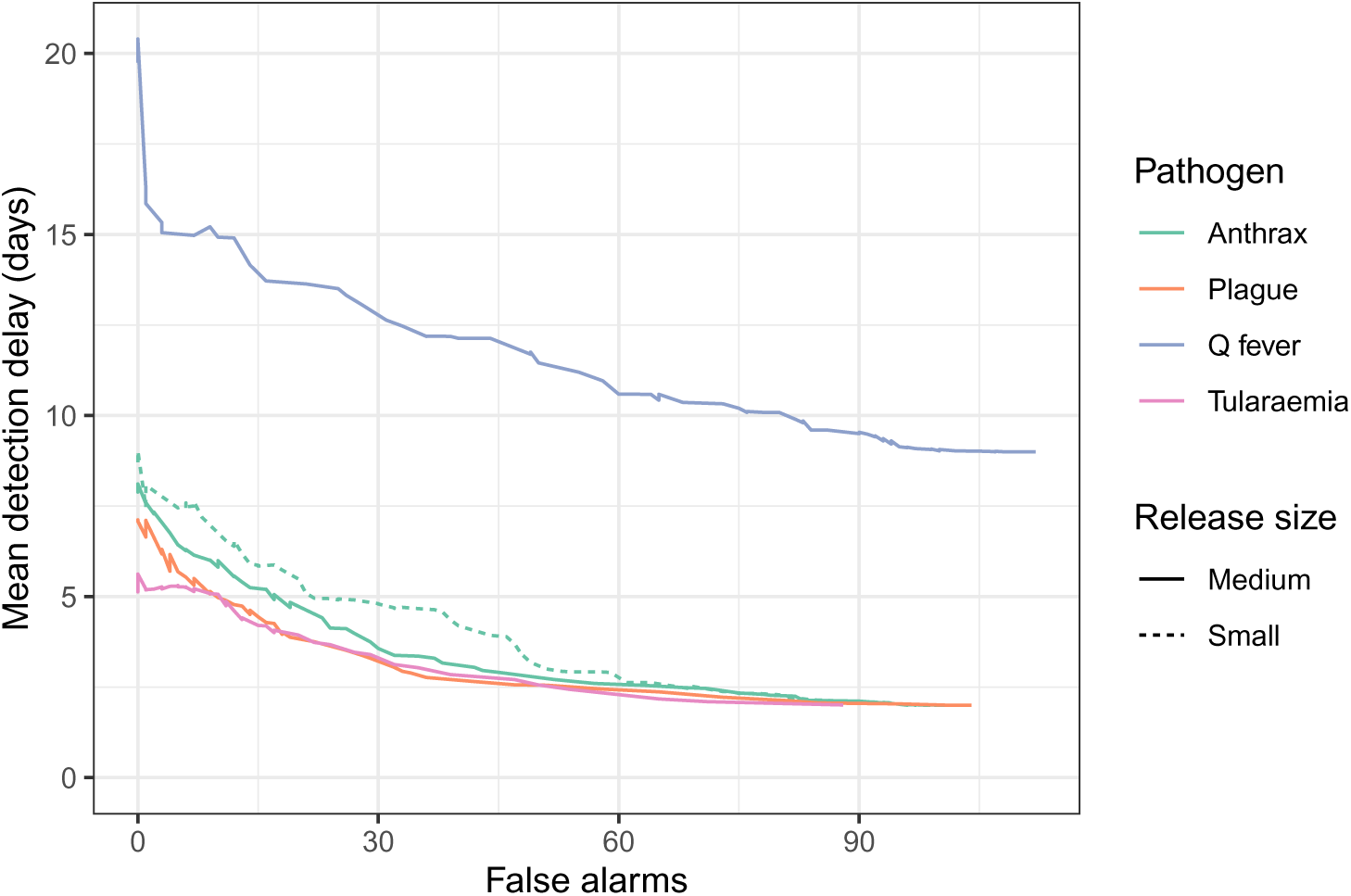
AMOC curves for mean delay from release to detection (study period of 113 days).

**Figure B10:**
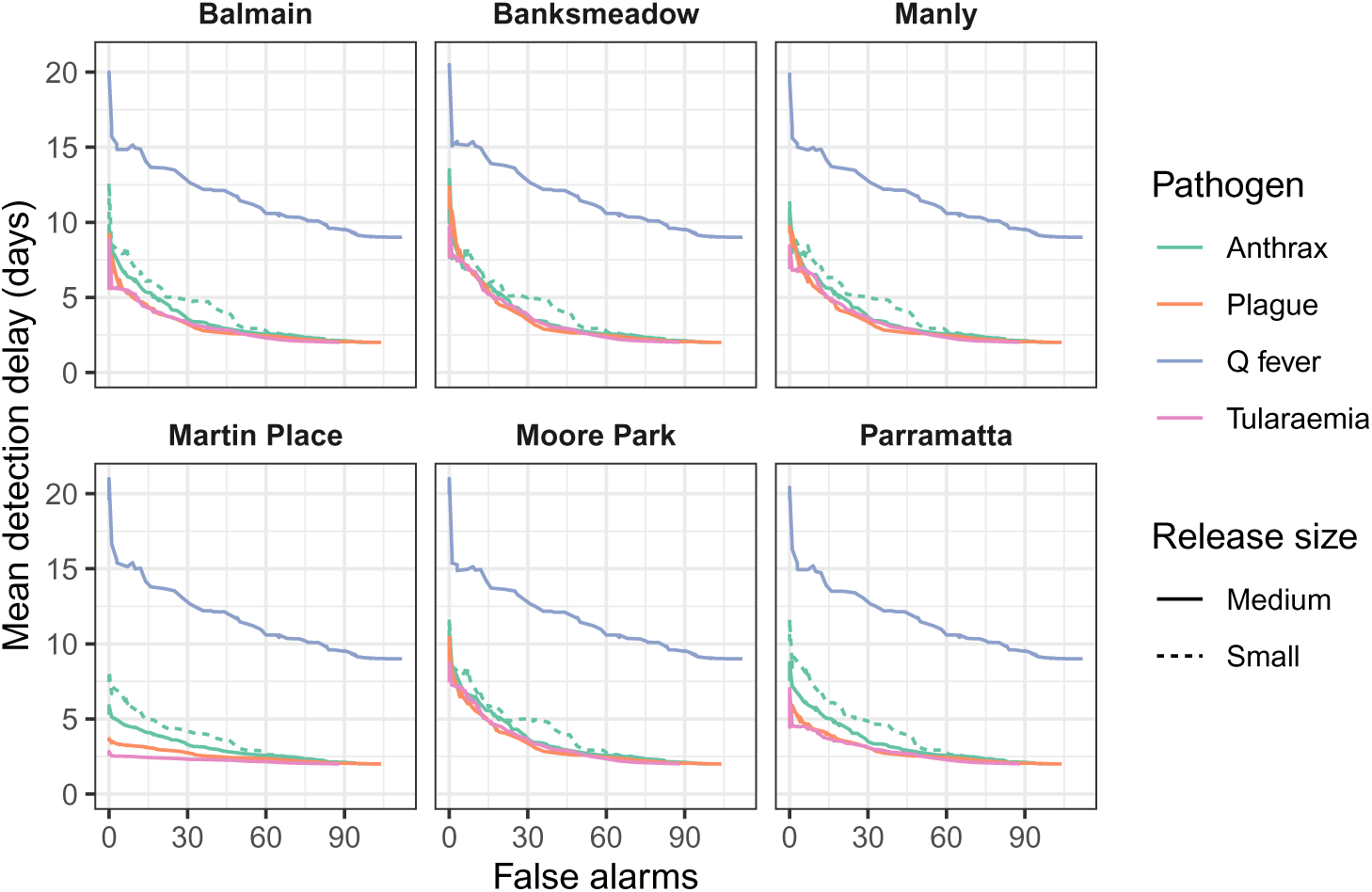
AMOC curves for mean delay from release to detection, shown separately for each release location (study period of 113 days).

Investigating the lead times separately for each release location (Figure B11) shows that anthrax, pneumonic plague, and tularaemia releases in Martin Place can be detected more quickly than the TTCD with few, if any, false alarms. In contrast, at very low false alarm rates, Q fever releases in Martin Place are detected with worse lead times than Q fever releases in other locations. This is due to the substantially larger number of exposures in Martin Place, due to the population density, which in turn results in earlier TTCDs with less variance than for other release locations (shown in Figure B3).

**Figure B11:**
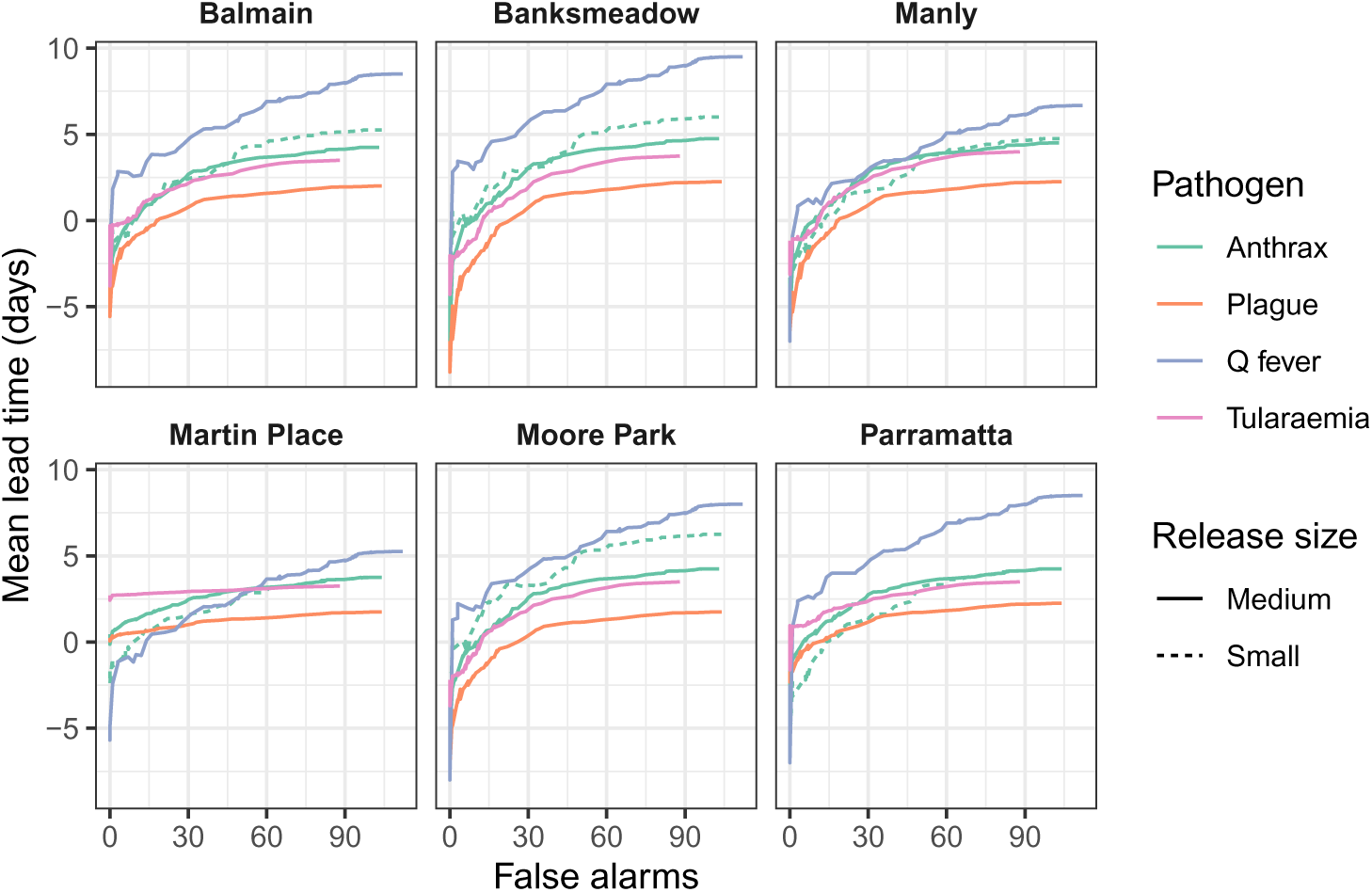
AMOC curves for mean lead time relative to clinical detection, shown separately for each release location (study period of 113 days).

Finally, we consider the trade-off between detection sensitivity and false alarm rates. Releases of anthrax, pneumonic plague, and tularaemia can be detected with very high sensitivity at low false alarm rates. In contrast, reliable detection of Q fever releases is only achieved at higher false alarm rates (e.g., 28 false alarms corresponds to one false alarm every 4 days). This is primarily due to the substantially smaller number of clinical events that occur in the Q fever releases. Consistent with detection timeliness, the highest sensitivity at low false alarm rates is achieved with higher population densities (Figure B12).

**Figure B12:**
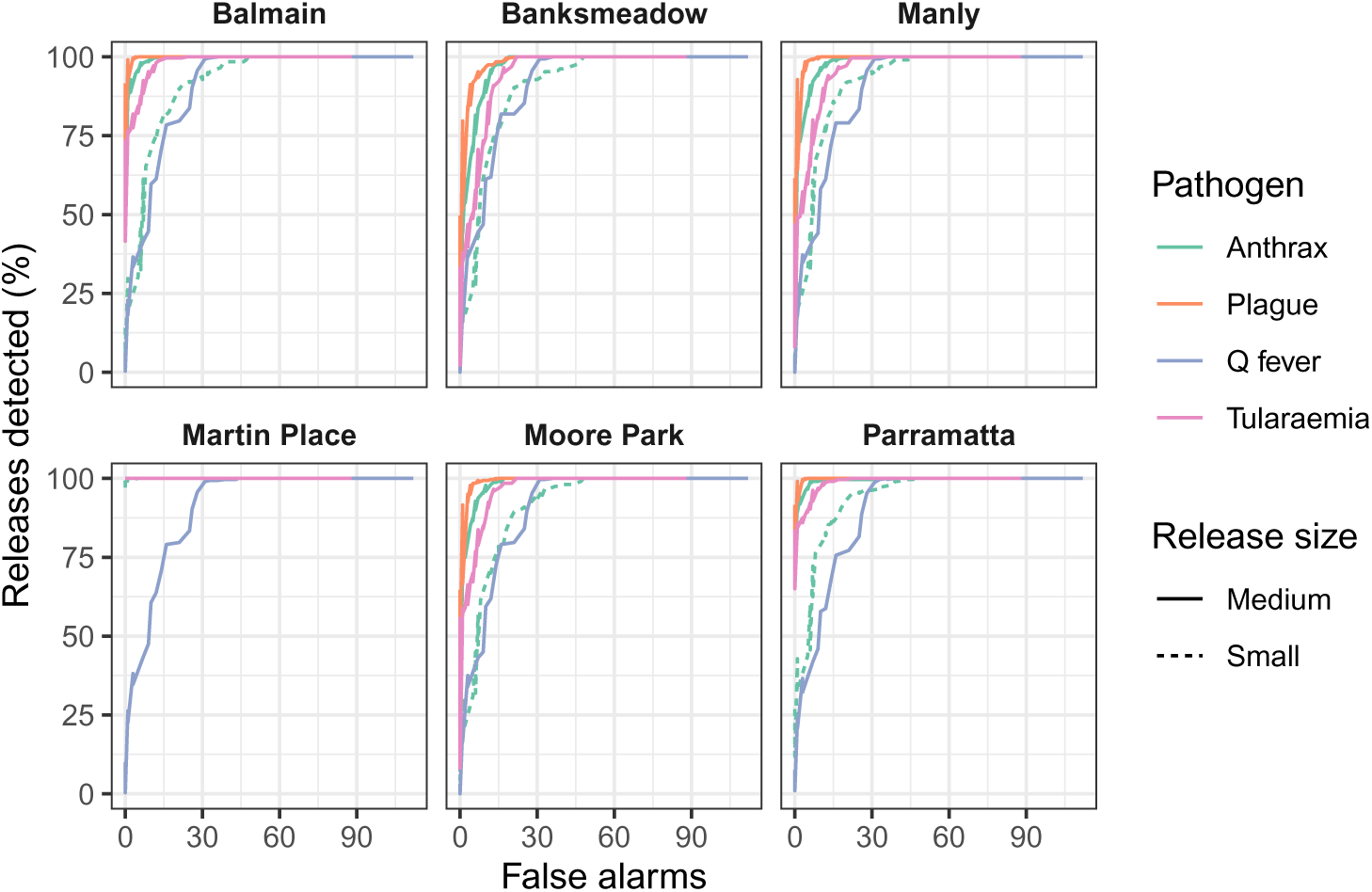
AMOC curves for the percentage of attacks that were detected by EpiDefend, shown separately for each release location (study period of 113 days).

